# Free water predicts dementia with Lewy bodies in isolated REM sleep behavior disorder

**DOI:** 10.1101/2025.05.15.25327709

**Authors:** Celine Haddad, Véronique Daneault, Violette Ayral, Marie Filiatrault, Alexandre Pastor-Bernier, Christina Tremblay, Arnaud Boré, Maxime Descoteaux, Andrew Vo, Jean-François Gagnon, Ronald B. Postuma, Petr Dusek, Stanislav Marecek, Zsoka Varga, Johannes Klein, Michele T. Hu, Stéphane Lehéricy, Isabelle Arnulf, Marie Vidailhet, Jean-Christophe Corvol, ICEBERG Study Group, Shady Rahayel

**Affiliations:** Center for Advanced Research in Sleep Medicine, Hôpital du Sacré-Cœur de Montréal – CIUSSS-NÎM, 5400 West Gouin Boulevard, Montreal H4J 1C5, Canada; Department of Psychology, University of Montreal, 90 Vincent d’Indy Avenue, Montreal H2V 2S9, Canada; Department of Neuroscience, University of Montreal, 2960 chemin de la Tour, Montreal H3T 1J4, Canada; Sherbrooke Connectivity Imaging Lab, University of Sherbrooke, 2500 Bd de l’Université, Sherbrooke J1K 0A5, Canada; The Neuro (Montreal Neurological Institute-Hospital), McGill University, 3801 University Street, Montreal H3A 2B4, Canada; Department of Psychology, Université du Québec à Montréal, 100 West Sherbrooke Street, Montreal H2X 3P2, Canada; Research Centre, Institut universitaire de gériatrie de Montréal, 4565 Queen Mary Rd, Montreal H3W 1W5, Canada; Department of Neurology, Montreal General Hospital, 1650 Cedar Avenue, Montreal H3G 1A4, Canada; Department of Neurology and Centre of Clinical Neurosciences, First Faculty of Medicine, Charles University and General University Hospital, Kateřinská 1660/32, 121 08 Nové Město, Prague, Czechia; Oxford Parkinson’s Disease Centre and Division of Neurology, Nuffield Department of Clinical Neurosciences, University of Oxford, Headly Way, Headington, OX3 9DU, Oxford, UK; Sorbonne Université, Institut du Cerveau – Paris Brain Institute – ICM, Assistance Publique Hôpitaux de Paris, INSERM 1127, CNRS 7225, Paris 75013, France; Department of Medicine, University of Montreal, 2900 Edouard Montpetit Boulevard, Montreal H3T 1J4, Canada

**Keywords:** REM sleep behavior disorder, Parkinson’s disease, dementia with Lewy bodies, free water, diffusion MRI, prognosis, biomarker

## Abstract

**INTRODUCTION:** Most individuals with isolated REM sleep behavior disorder (iRBD) develop dementia with Lewy bodies (DLB) or Parkinson’s disease (PD). Brain biomarkers predicting specific phenoconversion trajectories are lacking.

**METHODS:** In this multicenter diffusion MRI study (261 iRBD, 177 controls), free water (FW) was measured in the nucleus basalis of Meynert (NBM) and posterior substantia nigra (SN). Among 230 iRBD patients with follow-up, 64 converted (16 DLB, 38 PD). Time-to-event analyses were performed to assess differential phenoconversion.

**RESULTS:** Phenoconverters had higher FW in the NBM and posterior SN. Only FW in the NBM predicted conversion to DLB over PD. NBM volume predicted DLB conversion, but only FW remained significant when both were modeled. FW in the NBM correlated with lower MoCA scores in iRBD.

**DISCUSSION:** FW in the NBM is a sensitive biomarker of cognitive decline and DLB progression in iRBD, outperforming volume and supporting its use in early stratification.

## 1. Background

Isolated rapid eye movement (REM) sleep behavior disorder (iRBD) is a parasomnia characterized by the loss of normal muscle atonia during REM sleep, resulting in abnormal and often violent movements and vocalizations (1). It is widely recognized as an early manifestation of synucleinopathies such as dementia with Lewy bodies (DLB), Parkinson’s disease (PD), and multiple system atrophy (MSA) with over 90% of individuals progressing to one of these diseases within 15 years (2). Patients with iRBD experience brain changes similar to those seen in overt synucleinopathies, including brain atrophy (3-9), abnormal perfusion (10-14), and reduced striatal dopamine transporter binding (15-17). While existing imaging markers have been examined as potential predictors of disease progression, they have rarely been tested in large multicentric studies or across varying MRI parameters, and often lack specificity in distinguishing between progression to pathology such as DLB and PD (18). Given these limitations, there is a crucial need for novel biomarkers that can stratify iRBD patients based on neurodegeneration severity in targeted brain regions and predict differential progression to DLB or PD. This could guide early targeted neuroprotective interventions, potentially altering the course of the disease.

Free water (FW) quantification from diffusion MRI is a promising approach that quantifies extracellular isotropic diffusion FW to reflect neuroinflammation, edema, and neurodegeneration (19). This technique enhances traditional diffusion tensor imaging through a bi-compartment model that separates the isotropic diffusion of extracellular FW from the anisotropic, restricted diffusion within tissue (19). This approach has been applied in various neurodegenerative diseases to detect increases in FW, including PD, where increased FW content has been observed in the posterior region of the substantia nigra (SN) (20-26), a basal ganglia structure strongly associated with parkinsonism and synucleinopathies (27). FW increase in the SN has been associated with several disease markers and clinical features in PD, including reduced dopamine binding in the striatum (28) and motor features, particularly bradykinesia (29). FW increases in the basal forebrain (specifically in the nucleus basalis of Meynert (NBM)) of PD patients have also been reported in numerous studies, and have been associated with cognitive dysfunction, particularly attention and memory deficits (30-34). In DLB, similar FW alterations have been observed in the NBM and associated with cholinergic deficits and cognitive decline (35).

Although FW imaging has been studied in PD and DLB, there is limited data on its application in the prodromal phase such as iRBD. To date, only three single-center studies have investigated FW changes in the basal forebrain or posterior SN in iRBD patients (≤ 57 patients) compared to healthy controls (28, 31, 36). While one study did not find changes between iRBD and controls (36), the others found increased FW content, intermediate between those of controls and PD patients, in the basal forebrain or posterior SN (28, 31). This suggests that FW increases in iRBD reflect disease progression and early neurodegenerative changes. However, these studies were limited by the absence of longitudinal data and multicentric validation, raising uncertainties regarding the predictive value of FW for differential disease progression in iRBD. Understanding FW changes in the posterior SN and NBM may reveal markers of motor and cognitive pathways associated with phenoconversion from iRBD to PD or DLB.

In this study, we leveraged the largest collection of diffusion MRI scans from video-polysomnography (vPSG)-confirmed iRBD patients and healthy controls, obtained from ongoing prospective, longitudinal cohort studies across five sites worldwide. We processed the diffusion MRI data to extract FW content from the NBM and posterior SN and compared FW content between iRBD patients and controls. We assessed the clinical correlations of FW content in iRBD patients and investigated its predictive capability for determining specific disease pathology, including DLB and PD, or remaining disease-free. Finally, we investigated whether FW predicted phenoconversion independently from structural atrophy in the NBM. We hypothesized that increased FW content in the NBM and posterior SN would be predictive of specific phenoconversion in iRBD patients.

## 2. Methods

### 2.1. Sample

This longitudinal, multicenter prospective study included patients with vPSG-confirmed iRBD and healthy controls who underwent brain MRI, namely T1-weighted and diffusion-weighted scans. Recruitment was conducted independently at five sites within well-established longitudinal cohorts in sleep and movement disorders clinics. The sites included the Centre for Advanced Research on Sleep Medicine at the Hôpital du Sacré-Coeur de Montréal and The Neuro, Montreal, Canada; the First Faculty of Medicine at Charles University, Prague, Czechia; the Oxford Discovery Cohort, Oxford, UK; the Movement Disorders clinic at the Hôpital de la Pitié-Salpêtrière, Paris, France; and the Parkinson’s Progression Markers Initiative study (37).

iRBD was diagnosed according to the International Classification of Sleep Disorders (third edition) criteria, with all patients undergoing one night of vPSG recording (38). Following diagnosis, patients underwent neurological and cognitive assessments to confirm that RBD was in its isolated phase (iRBD). Patients were excluded if they had already developed an overt neurodegenerative synucleinopathy (DLB, PD or MSA) at the clinical evaluation closest to the MRI acquisition, as defined by established diagnostic criteria for DLB, PD, and MSA (39-41). Additional exclusion criteria included any history of brainstem stroke, a diagnosis of epilepsy, epileptiform abnormalities on EEG, antidepressant-triggered RBD, and RBD mimics such as sleepwalking, night terrors or untreated obstructive sleep apnea. All patients underwent the Montreal Cognitive Assessment (MoCA) to evaluate global cognitive function and the Movement Disorder Society-sponsored Unified Parkinson’s Disease Rating Scale (MDS-UPDRS-III) to assess the severity of parkinsonian motor features (42, 43). Patients were monitored prospectively with annual neurological and cognitive examinations to assess any conversion to PD, DLB or MSA. This study was approved by the research ethics committees at each participating site, with multicentric approval from the CIUSSS du Nord-de-l’Île-de-Montréal and the McGill University Health Centre. All procedures adhered to the ethical standards of the 1964 Declaration of Helsinki and its later amendments. A subset of these participants have been studied previously as part of multicentric imaging studies using T1- or diffusion-weighted MRI scans in iRBD (3, 4, 44, 45).

### 2.2. MRI Processing

T1- and diffusion-weighted brain MRI scans were acquired at each site using 3 Tesla MRI scanners, with acquisition parameters detailed in the Supplementary Material. All MRI scans were visually inspected for artifacts before processing (Figure S1 for workflow). The T1-weighted MRI scans were first processed using FreeSurfer (v.7.1.1), following established methods to segment the cortical and subcortical structures (3). The segmented maps and surfaces from FreeSurfer were then combined with the diffusion-weighted images and corresponding bval and bvec files as inputs to the TractoFlow-Atlas Based Segmentation (Tractoflow-ABS) pipeline (46) run on high-performance computing (HPC) resources. Tractoflow-ABS is an automated diffusion MRI processing pipeline designed to perform artefact removal, preprocessing, and tractography for robust, large-scale quantitative analyses (46). Key parameters for diffusion imaging processing included both single-shell and multi-shell data (when available, up to three shells: b = 0, 500, 1000, 2000). Free water estimation was performed using single-shell data, while multi-shell acquisitions were processed using appropriate multi-compartment models when available. Fiber response functions were set to [10, 3, 3], and tractography was conducted using local probabilistic tracking. Seeding was applied at the white matter/grey matter interface with 20 seeds per voxel. Spherical harmonics were set to order 6 for datasets with fewer than 32 gradient directions and to order 8 otherwise. FW fractions, ranging from 0 to 1 per voxel, were calculated using the FreeWater Flow tool (19, 47-49) and Python 3.8 with AMICO wheels on HPC resources. Key parameters included an axial diffusivity of 0.001 in the corpus callosum, a mean diffusivity of 0.0025 in the ventricles, and a radial diffusivity range from 0.0001 to 0.0065, with specific regularization settings.

To extract FW content, regions of interest (ROI) were aligned with the MNI152 template space. Basal forebrain ROIs were derived using a probabilistic atlas from postmortem data in the JuBrain Anatomy toolbox (version 2.2) with Mesulam’s nomenclature. Two distinct basal forebrain ROIs were selected, namely the left and right NBM, as the primary areas of interest. SN ROIs were defined using an isotropic 2-mm box in MarsBar, with coordinates set for left posterior SN (x = - 11, y = -21, z = -13) and right posterior SN (x = 11, y = -21, z = -13) as target areas, as well as left anterior SN (x = -9, y = -15, z = -13) and right anterior SN (x = 9, y = -15, z = -13) as additional control areas (Figure S1).

For mask registration, T1-weighted images were further processed using the CAT12 toolbox in SPM12 (release 6906) (50) under MATLAB R2018B to produce whole-brain warped and modulated maps per patient. This step involved segmentation into tissue classes, bias correction for intensity non-uniformity, and non-linear transformation to MNI template space. Transformation maps for aligning ROIs to patient space were generated using Advanced Normalization Tools with customized codes. This procedure involved registering each mask to the T1-warped image from Tractoflow using the antsApplyTransforms tool with the NearestNeighbor function. MarsBar-defined ROIs were similarly transformed. Customized scripts were then used to extract mean FW values for each ROI. FW values close to 0 indicate restricted water movement within tissue, while values near 1 reflect free diffusion in the extracellular space, indicating neuroinflammation.

To determine whether FW content in the basal forebrain predicted phenoconversion independently of regional atrophy, we also extracted gray matter volume from the same NBM regions in all participants. The NBM ROIs were identical to those used for FW extraction. Masks for the left and right NBM were co-registered and resliced to the MNI152 template using SPM12. Gray matter volume estimates (in mm^3^) were then extracted from unsmoothed, modulated gray matter maps using the “get_totals” script. Since volume scales with head size, total intracranial volume was extracted for each participant and all volume measures were normalized by dividing each regional volume by the corresponding total intracranial volume.

### 2.3. Statistical Analyses

To address variability in multicenter data collected using different scanners, the ComBat harmonization algorithm was applied to the FW values to correct for scanner-related effects (51, 52). This method has been validated in neuroimaging studies, including diffusion MRI (53), and enables robust site-level adjustment while preserving biological variability.

The primary outcome variable was phenoconversion status in iRBD patients, categorized into three groups: iRBD converters to DLB, iRBD converters to PD, and iRBD non-converters (disease-free at last follow-up). For cross-sectional analyses, group comparisons of normally distributed demographic, clinical, and imaging variables (FW and volume) were performed using independent samples t-tests for continuous variables and chi-squared tests for categorical variables. Non-normally distributed variables were assessed using the Mann-Whitney U test. Correlations between FW values in the NBM and posterior SN and clinical features (MoCA and MDS-UPDRS-III) were assessed using Pearson’s r, followed by partial correlations controlling for age and sex. Hemispheric differences in correlation strength were assessed using Hotelling’s t-test and Zou’s confidence interval method.

For longitudinal phenoconversion analyses, multinomial logistic regression was used to evaluate the predictive value of FW measures on phenoconversion odds. Model 1 tested FW in the NBM, Model 2 tested FW in the posterior SN, and Model 3 included both regions simultaneously, adjusting for age and sex in all models. To confirm regional specificity, these models were repeated using the FW values from the anterior SN as a control region.

To investigate time to phenoconversion, Kaplan-Meier survival analyses were performed. The MRI acquisition date was considered time 0, and follow-up time was computed as the interval to the latest clinical evaluation (or date of phenoconversion). Patients were stratified into high and low FW groups based on the median value within the iRBD sample. Log-rank tests were used to assess group differences in survival. Additional Kaplan-Meier curves were generated for specific conversion types (DLB versus PD versus non-converters). Cox proportional hazards regression models were used to assess whether FW content predicted time to phenoconversion, adjusting for age and sex. All FW values were z-scored to standardize interpretation of odds ratios and hazard ratios. Likelihood ratio tests were used to evaluate model fit. To examine whether FW predicted phenoconversion independently of structural atrophy, we also tested whether NBM volume predicted conversion type, and whether FW predicted phenoconversion when both FW and NBM volume were added in the same regression model. All statistical analyses were conducted using IBM SPSS Statistics (version 29.0) and RStudio (version 2023.06.2).

## 3. Results

### 3.1. Participants

The multicenter iRBD cohort initially included 592 participants, comprising 289 vPSG-confirmed iRBD patients and 303 controls, each with both T1- and diffusion-weighted brain MRI scans acquired across five centers as part of this prospective study (Figure 1). The cohort included 159 participants from Montreal, 140 from Prague, 132 from Oxford, 107 from Paris, and 54 from the Parkinson’s Progression Markers Initiative study. All scans successfully passed initial diffusion MRI processing; however, 25 were excluded during visual quality control due to misregistration, resulting in 567 scans eligible for analysis. To ensure comparability between iRBD and control groups, 129 scans were excluded for statistical matching on age and sex. While each iRBD site included matched controls, the PPMI controls included a higher proportion of younger females. Most exclusions therefore occurred in the control group, with 126 controls and 28 iRBD patients excluded. All MRI processing and FW estimation were performed prior to matching to prevent post-hoc quality control from introducing imbalances. A total of 438 participants (261 iRBD patients, 177 controls) were therefore retained for analyses involving the posterior SN, with no further exclusions, as all scans showed satisfactory alignment.

**Figure 1.**
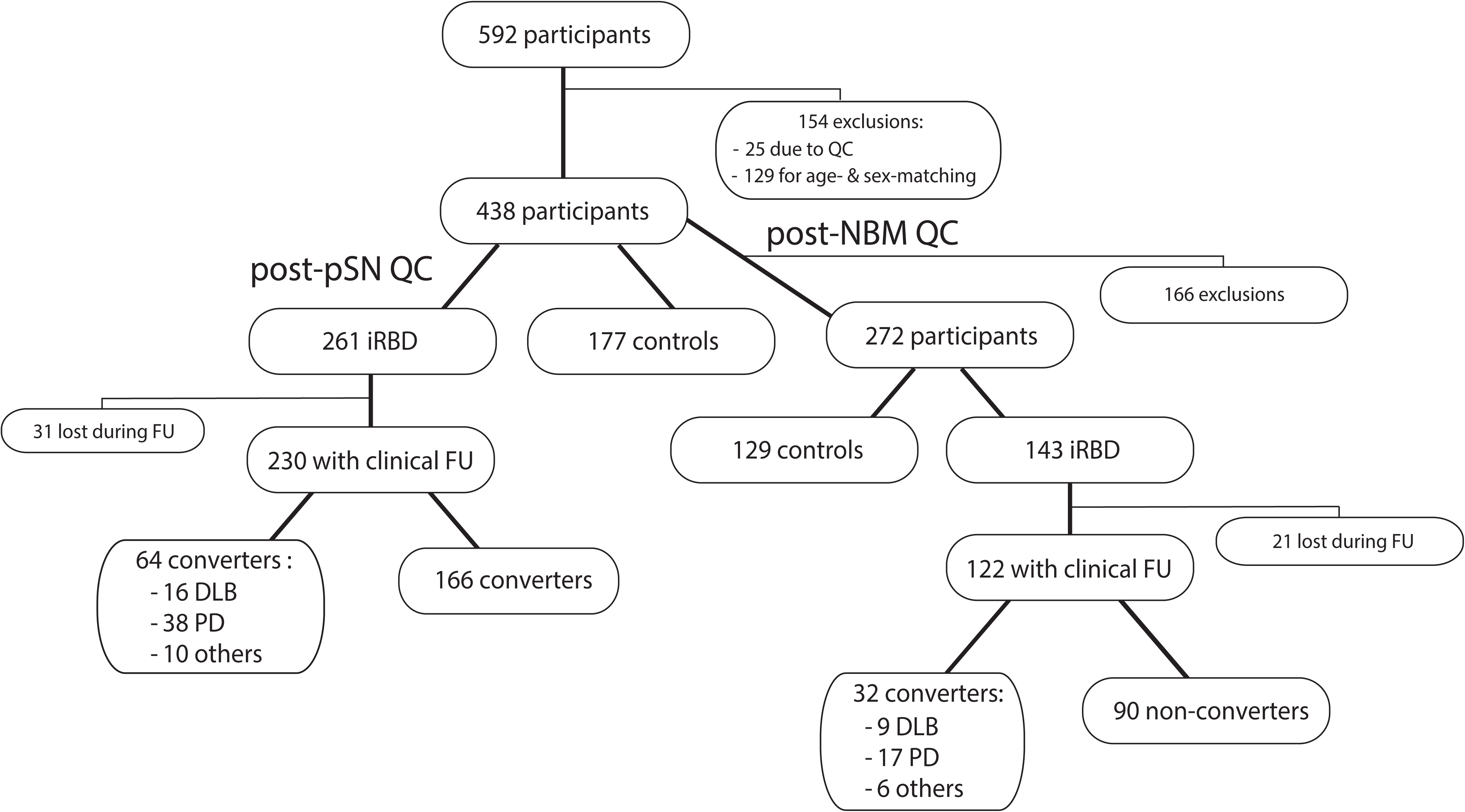
Flowchart of participant selection. Flowchart illustrating participant selection and inclusion for NBM and pSN analyses. Participants were excluded based on quality control, demographic matching, and availability of clinical measures. DLB = dementia with Lewy bodies; FW = free water; iRBD = isolated REM sleep behavior disorder; NBM = nucleus basalis of Meynert; PD = Parkinson’s disease; pSN = posterior substantia nigra; QC = quality control.

In this sample, iRBD patients had a mean age of 66.3 years (SD: 6.5), with 32 (12%) identifying as female. The mean MoCA score was 25.4 (SD: 3.0) and the mean score on the MDS-UPDRS-III was 6.4 (SD: 5.5) (Table 1). iRBD patients had significantly lower MoCA scores and higher MDS-UPDRS-III scores compared to controls (P < 0.001). As expected based on the initial group compositions, demographic comparisons between included and excluded participants reflected the effects of matching. Excluded controls were significantly younger (58.1 years, P < 0.001) and more often female (72%, P < 0.001), while excluded iRBD patients were older (73.9 years, P < 0.001). No significant differences were observed in MoCA scores (P = 0.24), MDS-UPDRS-III (P = 0.91), or sex (P = 0.20) among excluded versus included iRBD patients. Following a strict quality control procedure for the NBM due to its anatomical proximity to CSF, which can contaminate FW estimates, 272 MRI scans were retained for this region, comprising 143 iRBD patients and 129 controls with similar age (P = 0.62) and sex distribution (P = 0.13). Patients with iRBD included in the NBM (N = 143) and posterior SN analyses (N = 261) did not differ in age (P = 0.33), sex distribution (P = 0.73), MoCA scores (P = 0.40), and MDS-UPDRS-III scores (P = 0.50).

**Table 1.**
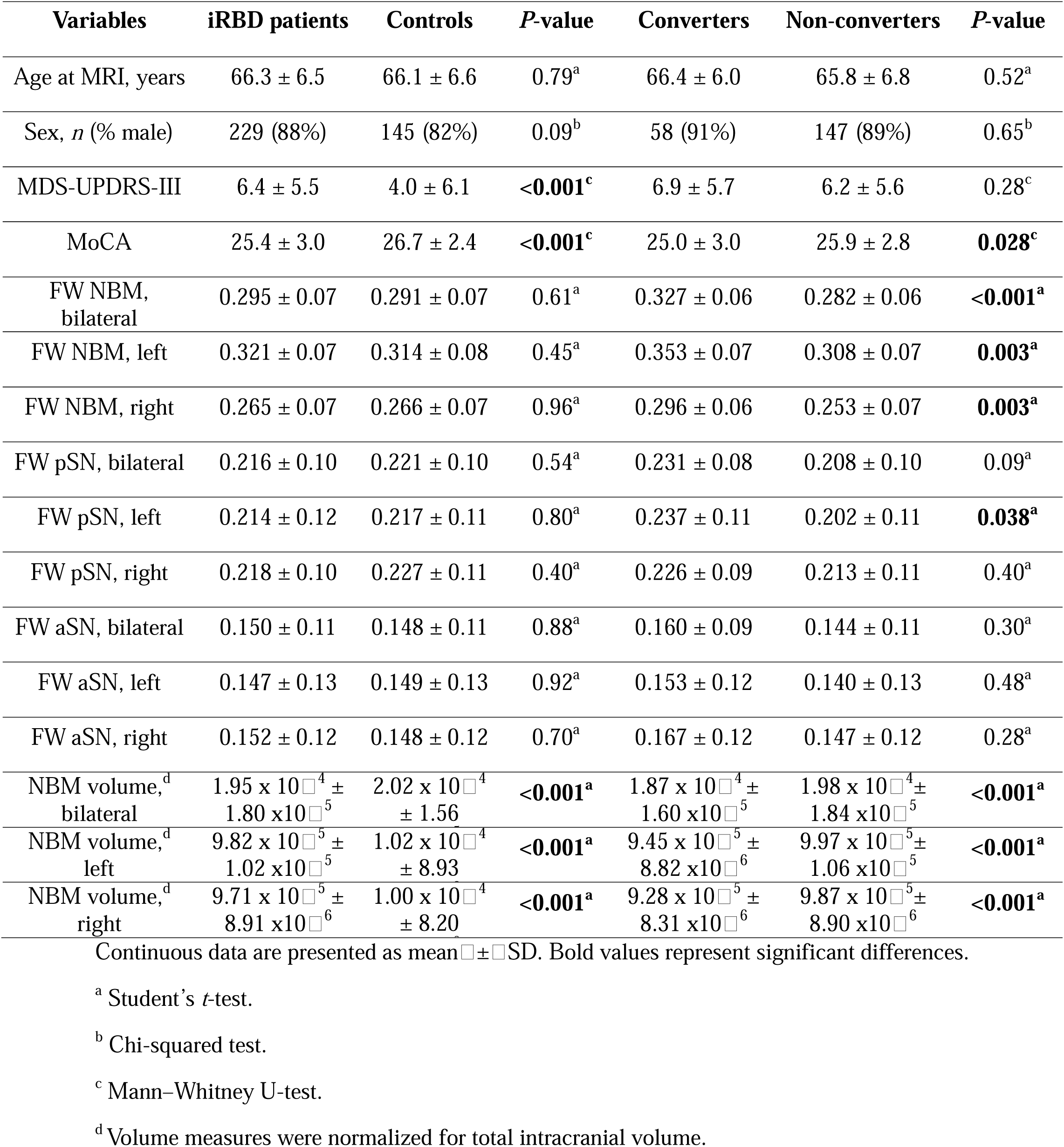
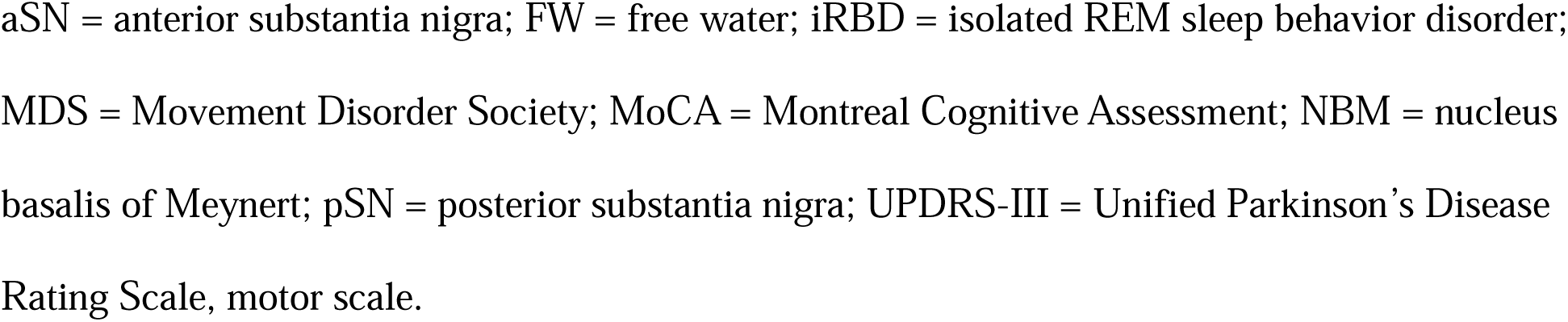
Demographic, clinical, and imaging variables of iRBD patients and controls.

| Variables | iRBD patients | Controls | *P*-value | Converters | Non-converters | *P*-value |
| --- | --- | --- | --- | --- | --- | --- |
| Age at MRI, years | 66.3 ± 6.5 | 66.1 ± 6.6 | 0.79<sup>a</sup> | 66.4 ± 6.0 | 65.8 ± 6.8 | 0.52<sup>a</sup> |
| Sex, *n* (% male) | 229 (88%) | 145 (82%) | 0.09<sup>b</sup> | 58 (91%) | 147 (89%) | 0.65<sup>b</sup> |
| MDS-UPDRS-III | 6.4 ± 5.5 | 4.0 ± 6.1 | **<0.001<sup>c</sup>** | 6.9 ± 5.7 | 6.2 ± 5.6 | 0.28<sup>c</sup> |
| MoCA | 25.4 ± 3.0 | 26.7 ± 2.4 | **<0.001<sup>c</sup>** | 25.0 ± 3.0 | 25.9 ± 2.8 | **0.028<sup>c</sup>** |
| FW NBM, bilateral | 0.295 ± 0.07 | 0.291 ± 0.07 | 0.61<sup>a</sup> | 0.327 ± 0.06 | 0.282 ± 0.06 | **<0.001<sup>a</sup>** |
| FW NBM, left | 0.321 ± 0.07 | 0.314 ± 0.08 | 0.45<sup>a</sup> | 0.353 ± 0.07 | 0.308 ± 0.07 | **0.003<sup>a</sup>** |
| FW NBM, right | 0.265 ± 0.07 | 0.266 ± 0.07 | 0.96<sup>a</sup> | 0.296 ± 0.06 | 0.253 ± 0.07 | **0.003<sup>a</sup>** |
| FW pSN, bilateral | 0.216 ± 0.10 | 0.221 ± 0.10 | 0.54<sup>a</sup> | 0.231 ± 0.08 | 0.208 ± 0.10 | 0.09<sup>a</sup> |
| FW pSN, left | 0.214 ± 0.12 | 0.217 ± 0.11 | 0.80<sup>a</sup> | 0.237 ± 0.11 | 0.202 ± 0.11 | **0.038<sup>a</sup>** |
| FW pSN, right | 0.218 ± 0.10 | 0.227 ± 0.11 | 0.40<sup>a</sup> | 0.226 ± 0.09 | 0.213 ± 0.11 | 0.40<sup>a</sup> |
| FW aSN, bilateral | 0.150 ± 0.11 | 0.148 ± 0.11 | 0.88<sup>a</sup> | 0.160 ± 0.09 | 0.144 ± 0.11 | 0.30<sup>a</sup> |
| FW aSN, left | 0.147 ± 0.13 | 0.149 ± 0.13 | 0.92<sup>a</sup> | 0.153 ± 0.12 | 0.140 ± 0.13 | 0.48<sup>a</sup> |
| FW aSN, right | 0.152 ± 0.12 | 0.148 ± 0.12 | 0.70<sup>a</sup> | 0.167 ± 0.12 | 0.147 ± 0.12 | 0.28<sup>a</sup> |
| NBM volume,<sup>d</sup> bilateral | 1.95 x 10<sup>-4</sup> ± 1.80 x10<sup>-5</sup> | 2.02 x 10<sup>-4</sup> ± 1.56 | **<0.001<sup>a</sup>** | 1.87 x 10<sup>-4</sup> ± 1.60 x10<sup>-5</sup> | 1.98 x 10<sup>-4</sup> ± 1.84 x10<sup>-5</sup> | **<0.001<sup>a</sup>** |
| NBM volume,<sup>d</sup> left | 9.82 x 10<sup>-5</sup> ± 1.02 x10<sup>-5</sup> | 1.02 x 10<sup>-4</sup> ± 8.93 | **<0.001<sup>a</sup>** | 9.45 x 10<sup>-5</sup> ± 8.82 x10<sup>-6</sup> | 9.97 x 10<sup>-5</sup> ± 1.06 x10<sup>-5</sup> | **<0.001<sup>a</sup>** |
| NBM volume,<sup>d</sup> right | 9.71 x 10<sup>-5</sup> ± 8.91 x10<sup>-6</sup> | 1.00 x 10<sup>-4</sup> ± 8.20 | **<0.001<sup>a</sup>** | 9.28 x 10<sup>-5</sup> ± 8.31 x10<sup>-6</sup> | 9.87 x 10<sup>-5</sup> ± 8.90 x10<sup>-6</sup> | **<0.001<sup>a</sup>** |

Continuous data are presented as mean ± SD. Bold values represent significant differences.
<sup>a</sup> Student's *t*-test.
<sup>b</sup> Chi-squared test.
<sup>c</sup> Mann–Whitney U-test.
<sup>d</sup> Volume measures were normalized for total intracranial volume.
aSN = anterior substantia nigra; FW = free water; iRBD = isolated REM sleep behavior disorder; MDS = Movement Disorder Society; MoCA = Montreal Cognitive Assessment; NBM = nucleus basalis of Meynert; pSN = posterior substantia nigra; UPDRS-III = Unified Parkinson's Disease Rating Scale, motor scale.

### 3.2. Free water does not distinguish between iRBD patients and controls

We first investigated whether FW content in the NBM and posterior SN differed between the overall group of iRBD patients and controls. In the NBM, the mean FW value was 0.295 (SD: 0.07) for patients and 0.291 (SD: 0.07) for controls, with a non-significant mean difference of -0.004 (95% CI: -0.02 to 0.01; P= 0.61) (Figure 2 and Table 1). Similarly, in the bilateral posterior SN, the mean FW value was 0.216 (SD: 0.10) for patients and 0.221 (SD: 0.10) for controls, with a non-significant mean difference of 0.006 (95% CI: -0.01 to 0.02; P = 0.54) (Figure 2 and Table 1). Furthermore, no significant differences in FW content was observed between groups when the NBM and posterior SN were separated into left and right hemispheres or when analyzing the anterior SN as a control region (Figure 2, Table 1 and Figure S2). Controls excluded post-processing and matching had lower FW in the NBM (0.24, P < 0.001), while excluded iRBD patients had higher NBM FW (0.37, P = 0.013), with no significant differences in the posterior SN or anterior SN (P > 0.05).

**Figure 2.**
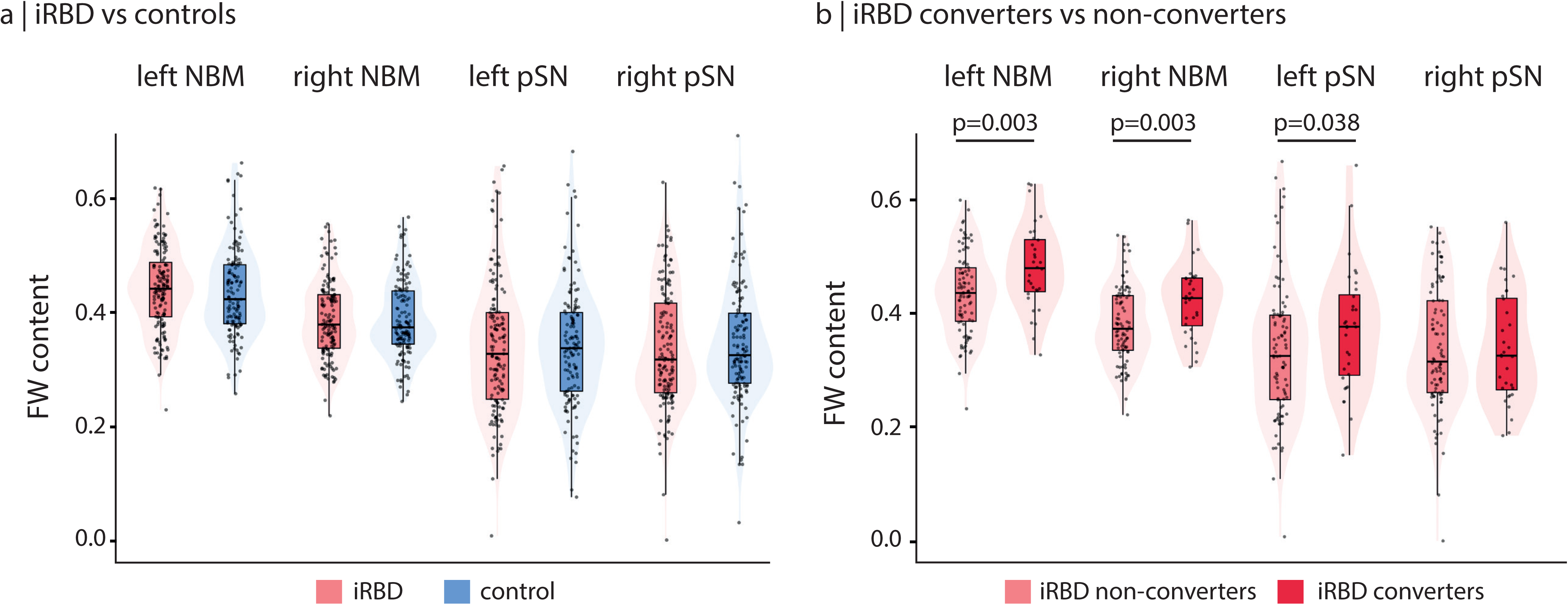
FW levels in iRBD patients and controls. a) Box plots and violin plots showing the distribution of FW content in the NBM and pSN for iRBD patients (red) and healthy controls (blue). FW content is presented after harmonization for scanner effects, with values standardized to positive values for visualization purposes. No significant differences were observed between groups in either region. b) Box plots and violin plots showing the distribution of FW content in the NBM and posterior SN for iRBD converters (dark red) and non-converters (light red). FW content in the NBM and pSN were significantly increased in iRBD converters compared to non-converters. FW = free water; iRBD = isolated REM sleep behavior disorder; NBM = nucleus basalis of Meynert; pSN = posterior substantia nigra.

### 3.3. Free water in the NBM is associated with cognitive decline

We examined whether FW content in the NBM and posterior SN were correlated with MoCA and MDS-UPDRS-III scores in iRBD patients. We observed a significant negative correlation between FW in the NBM and MoCA scores in iRBD patients, with increased FW content associated with decreased MoCA scores (r = -0.25, P = 0.004) (Figure 3). This association was significant in both hemispheres, with a correlation of r = -0.27 (P = 0.002) in the left hemisphere and r = -0.18 (P = 0.036) in the right hemisphere. The strength of the correlation between FW and MoCA did not significantly differ between hemispheres (coefficient difference = -0.08, 95% CI: -0.23 to 0.06, P = 0.23). However, after adjusting for age and sex, the association remained significant only for the left NBM. In contrast, there were no significant correlations between FW content in either the NBM or the posterior SN and MDS-UPDRS-III scores in iRBD patients (P = 0.45 for NBM, P = 0.58 for posterior SN). Additionally, FW content in the posterior SN did not show a significant correlation with MoCA scores in iRBD patients (P = 0.59), nor were there significant correlations with FW in the control group (all P > 0.05).

**Figure 3.**
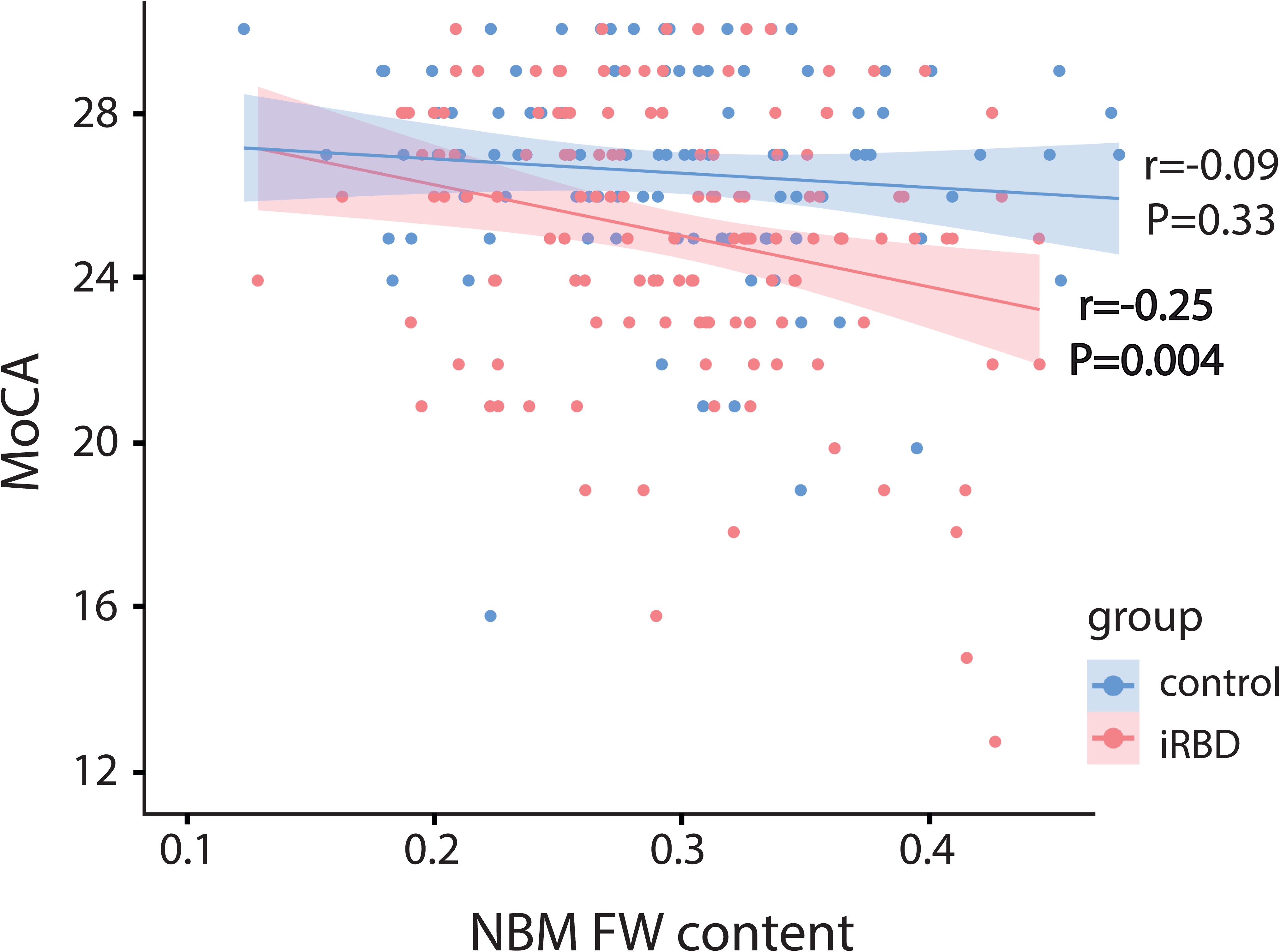
Association between FW levels in the NBM and MoCA scores. Scatter plot showing the relationship between FW content in the NBM and MoCA scores in iRBD patients (red) and healthy controls (blue). The shaded areas represent the 95% confidence intervals for the regression lines. In iRBD, higher FW content in the NBM was significantly associated with lower MoCA scores; no significant correlation was observed in controls. FW = free water; iRBD = isolated REM sleep behavior disorder; MoCA = Montreal Cognitive Assessment; NBM = nucleus basalis of Meynert.

### 3.4. Increased FW is associated with higher risk of phenoconversion in iRBD

To assess whether FW may be associated with disease progression in iRBD, we compared FW content in the NBM and posterior SN between iRBD patients who progressed to an overt synucleinopathy during follow-up versus those who remained disease-free. Of the 261 patients, 31 (12%) were lost to follow-up, resulting in a sample of 230 iRBD patients followed longitudinally. There were no significant differences in FW values between patients lost to follow-up and those followed longitudinally for the NBM (P = 0.56), posterior SN (P = 0.45), anterior SN (P = 0.55) nor in age (P = 0.06), sex (p = 0.06), and MDS-UPDRS-III score (P = 0.38). However, a significant difference was observed in MoCA scores (P = 0.002), with patients lost to follow-up showing lower cognitive performance compared to those retained (23.7 versus 25.7, p = 0.002). Patients were followed for a median of 8.4 years (range: 4 months to 16 years). At their latest clinical examination, 64 (28%) patients had converted to a neurodegenerative disease (converters), while 166 (72%) remained disease-free (non-converters). There were no significant differences between converters and non-converters regarding age at MRI scan (P = 0.52), sex (P = 0.65), and MDS-UPDRS-III score (P = 0.28) (Table 1). However, iRBD converters had lower scores on the MoCA (P = 0.028) (Table 1).

When stratifying iRBD patients by phenoconversion status, we found significant differences in FW content between converters and non-converters. iRBD converters exhibited higher FW values bilaterally in the NBM compared to non-converters, with a mean difference of -0.044 ± 0.013 (95% CI: -0.070 to -0.019, P < 0.001) (Table 1 and Figure 2). This increase was significant in both the left (mean difference: -0.045 ± 0.015, 95% CI: -0.074 to -0.016, P = 0.003) and right NBM (mean difference: -0.042 ± 0.038, 95% CI: -0.069 to -0.015, P = 0.003) (Table 1). In the posterior SN, iRBD converters showed increased FW only on the left side (mean difference: -0.034 ± 0.017, 95% CI: -0.068 to -0.002, P = 0.038) (Table 1 and Figure 2). To verify the specificity of these findings, we further assessed FW content in the anterior SN as a control region and did not find significant changes between groups (P = 0.48 on the left and P = 0.28 on the right) (Table 1 and Figure S3). When comparing iRBD subgroups to controls, we found that iRBD non-converters had similar FW content in the NBM (P = 0.34) and posterior SN (P = 0.23) compared to controls.

Of the 230 iRBD patients with follow-up, 67 had missing clinical information for time-to-event analyses, resulting in a post-quality-controlled sample of 163 (48 converted) for posterior SN analyses and 86 (24 converted) for NBM analyses. Kaplan-Meier survival analyses were conducted to assess whether FW content in the NBM and posterior SN predicted time to phenoconversion (in years) in iRBD. Patients were dichotomized into high and low FW groups based on the median FW content across the iRBD cohort. Survival curves revealed that higher FW content in both the NBM (P = 0.016) or posterior SN (P = 0.00084) was associated with a significantly increased risk of phenoconversion over time (Figure 4). When adjusting for age and sex using Cox proportional hazards regression models, increased FW in both regions remained significant predictors of phenoconversion. Specifically, higher FW in the NBM was associated with a two-fold increased risk of conversion (HR = 2.35, 95% CI: 1.36 to 4.05, P = 0.002) and increased FW in the posterior SN was linked to a 1.3-fold increased conversion risk (HR = 1.33, 95% CI: 1.00-1.77, P = 0.048) (Table S1). Among patients with high FW content in the NBM, 80% remained disease-free at 2 years compared to 95% of those with low FW content; survival dropped to 68% at 4 years and 60% at 8 years (Figure 4A). Similarly, for the posterior SN, 79% of patients with high FW content were disease-free at 2 years (versus 94% with low FW), 67% at 4 years, and 58% at 8 years (Figure 4B). However, importantly, Cox regression models including FW in both the NBM and posterior SN, along with age and sex, revealed that only NBM FW was a significant predictor of phenoconversion in iRBD (HR = 2.30, 95% CI: 1.33 to 3.98, P = 0.003), and not posterior SN (HR = 1.12, 95% CI: 0.74 to 1.69, P = 0.58) (Table S1). This indicates that while increased FW content in the NBM or posterior SN are associated with a higher risk of phenoconversion in iRBD, only FW in the NBM predicts this risk when considered simultaneously.

**Figure 4.**
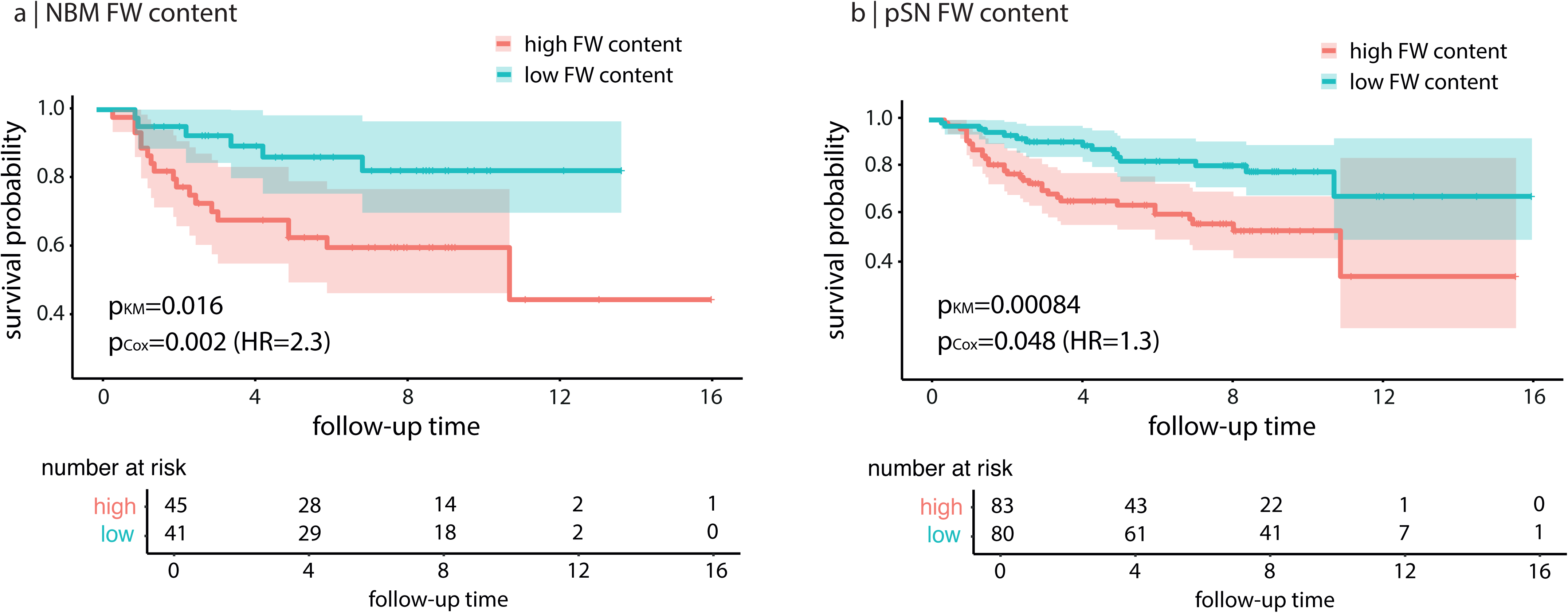
Survival analysis of phenoconversion risk in iRBD based on FW content. Kaplan-Meier survival curves showing survival probabilities (i.e., remaining disease-free at the latest clinical follow-up compared to receiving a diagnosis of neurodegenerative disease) over time based on FW content in **(a)** the NBM and **(b)** the pSN. High FW content based on the median FW level is represented in red and low FW content is represented in blue. Shaded areas indicate the 95% confidence intervals for each group. FW = free water; iRBD = isolated REM sleep behavior disorder; NBM = nucleus basalis of Meynert; pSN = posterior substantia nigra.

### 3.5. Free water in the NBM predicts DLB over PD in iRBD

We next investigated whether FW content in the NBM and posterior SN could predict specific phenoconversion pathways (i.e., DLB versus PD) in iRBD. Among the 230 iRBD patients with longitudinal follow-up, 10 (4%) converted to other neurodegenerative syndromes (e.g., MSA or pure autonomic failure) and were excluded, yielding a sample of 220 iRBD patients. These were classified as converters to DLB (N = 16), converters to PD (N = 38) or non-converters (N = 166). Model 1, testing FW in the NBM while adjusting for age and sex, revealed that higher FW content significantly predicted conversion to DLB compared to remaining disease-free (B = 1.94, P < 0.001), with an OR of 6.92 (95% CI: 2.28 to 21.04) per one-SD increase in NBM FW (Table 2). In contrast, NBM FW did not significantly predict conversion to PD (B = 0.51, P = 0.11, OR = 1.67, 95% CI: 0.89 to 3.15). However, FW in the NBM did differentiate DLB from PD converters (B = 1.42, P = 0.019), with an OR of 4.14 (95% CI: 1.26 to 13.55), suggesting selective sensitivity to DLB progression (Table 2).

**Table 2.**
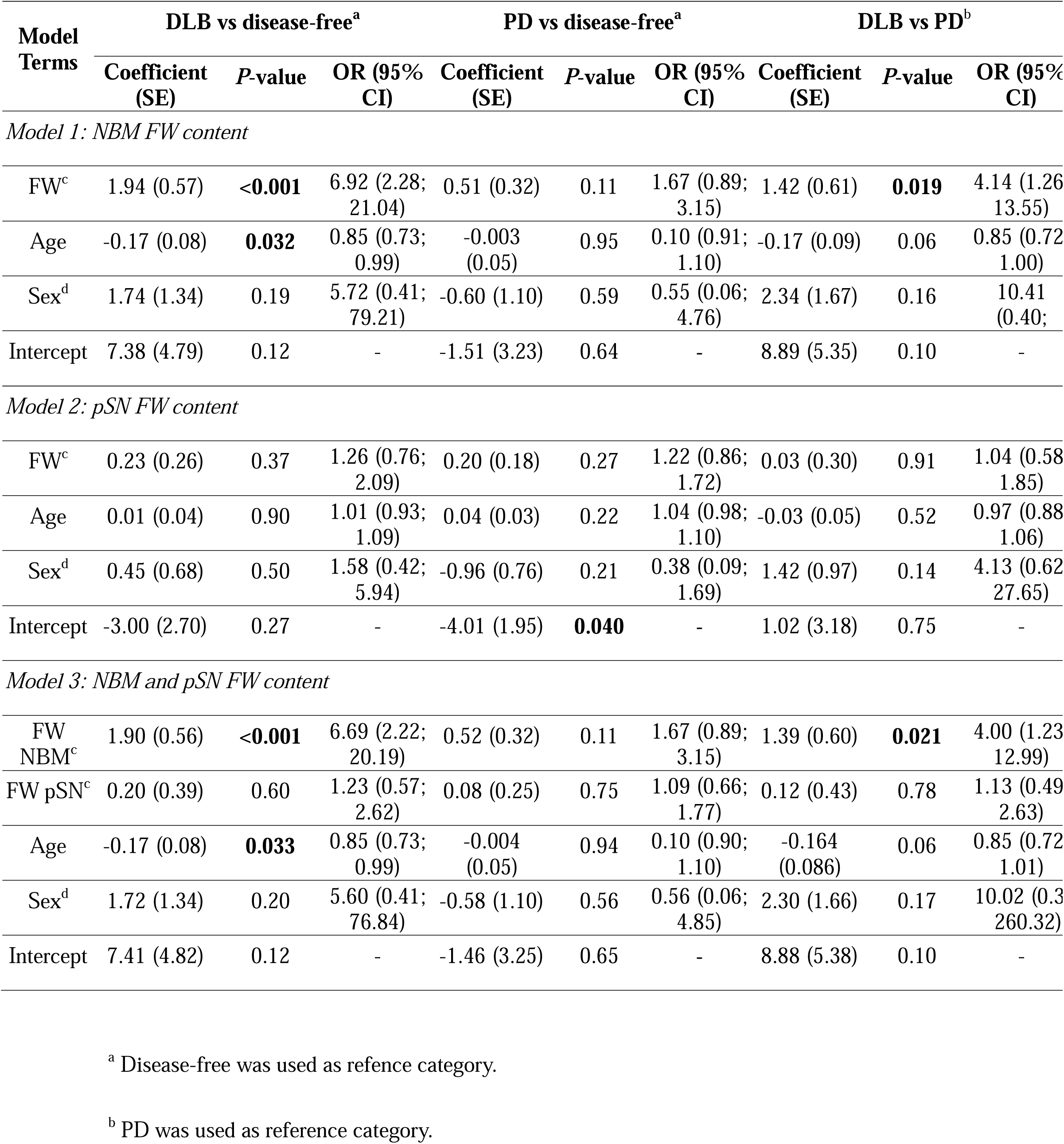
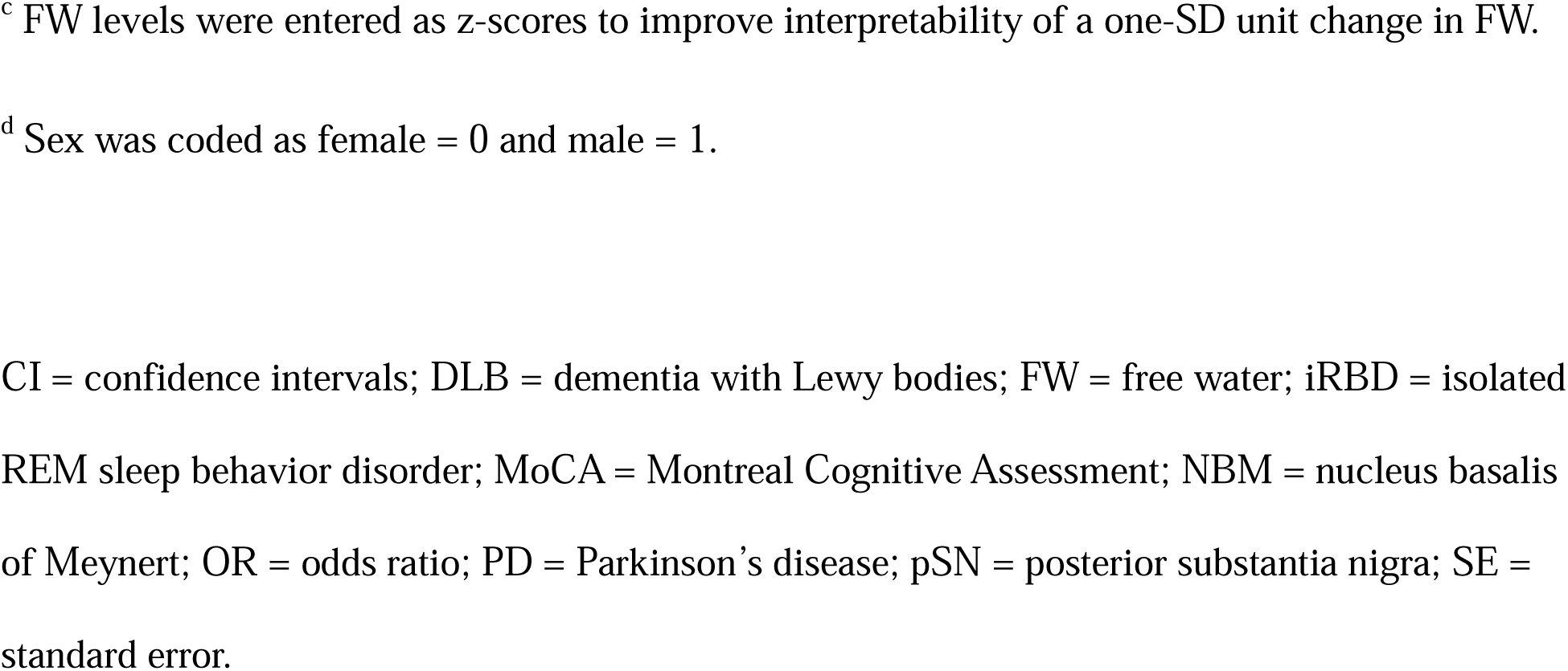
Logistic regression models of FW predicting phenoconversion in iRBD.

| Model Terms | DLB vs disease-free<sup>a</sup> |  |  | PD vs disease-free<sup>a</sup> |  |  | DLB vs PD<sup>b</sup> |  |  |
| --- | --- | --- | --- | --- | --- | --- | --- | --- | --- |
|  | Coefficient (SE) | P-value | OR (95% CI) | Coefficient (SE) | P-value | OR (95% CI) | Coefficient (SE) | P-value | OR (95% CI) |
| *Model 1: NBM FW content* |  |  |  |  |  |  |  |  |  |
| FW<sup>c</sup> | 1.94 (0.57) | **<0.001** | 6.92 (2.28; 21.04) | 0.51 (0.32) | 0.11 | 1.67 (0.89; 3.15) | 1.42 (0.61) | **0.019** | 4.14 (1.26 13.55) |
| Age | -0.17 (0.08) | **0.032** | 0.85 (0.73; 0.99) | -0.003 (0.05) | 0.95 | 0.10 (0.91; 1.10) | -0.17 (0.09) | 0.06 | 0.85 (0.72 1.00) |
| Sex<sup>d</sup> | 1.74 (1.34) | 0.19 | 5.72 (0.41; 79.21) | -0.60 (1.10) | 0.59 | 0.55 (0.06; 4.76) | 2.34 (1.67) | 0.16 | 10.41 (0.40; ) |
| Intercept | 7.38 (4.79) | 0.12 | - | -1.51 (3.23) | 0.64 | - | 8.89 (5.35) | 0.10 | - |
| *Model 2: pSN FW content* |  |  |  |  |  |  |  |  |  |
| FW<sup>c</sup> | 0.23 (0.26) | 0.37 | 1.26 (0.76; 2.09) | 0.20 (0.18) | 0.27 | 1.22 (0.86; 1.72) | 0.03 (0.30) | 0.91 | 1.04 (0.58 1.85) |
| Age | 0.01 (0.04) | 0.90 | 1.01 (0.93; 1.09) | 0.04 (0.03) | 0.22 | 1.04 (0.98; 1.10) | -0.03 (0.05) | 0.52 | 0.97 (0.88 1.06) |
| Sex<sup>d</sup> | 0.45 (0.68) | 0.50 | 1.58 (0.42; 5.94) | -0.96 (0.76) | 0.21 | 0.38 (0.09; 1.69) | 1.42 (0.97) | 0.14 | 4.13 (0.62 27.65) |
| Intercept | -3.00 (2.70) | 0.27 | - | -4.01 (1.95) | **0.040** | - | 1.02 (3.18) | 0.75 | - |
| *Model 3: NBM and pSN FW content* |  |  |  |  |  |  |  |  |  |
| FW NBM<sup>c</sup> | 1.90 (0.56) | **<0.001** | 6.69 (2.22; 20.19) | 0.52 (0.32) | 0.11 | 1.67 (0.89; 3.15) | 1.39 (0.60) | **0.021** | 4.00 (1.23 12.99) |
| FW pSN<sup>c</sup> | 0.20 (0.39) | 0.60 | 1.23 (0.57; 2.62) | 0.08 (0.25) | 0.75 | 1.09 (0.66; 1.77) | 0.12 (0.43) | 0.78 | 1.13 (0.49 2.63) |
| Age | -0.17 (0.08) | **0.033** | 0.85 (0.73; 0.99) | -0.004 (0.05) | 0.94 | 0.10 (0.90; 1.10) | -0.164 (0.086) | 0.06 | 0.85 (0.72 1.01) |
| Sex<sup>d</sup> | 1.72 (1.34) | 0.20 | 5.60 (0.41; 76.84) | -0.58 (1.10) | 0.56 | 0.56 (0.06; 4.85) | 2.30 (1.66) | 0.17 | 10.02 (0.3 260.32) |
| Intercept | 7.41 (4.82) | 0.12 | - | -1.46 (3.25) | 0.65 | - | 8.88 (5.38) | 0.10 | - |

<sup>a</sup> Disease-free was used as reference category.
<sup>b</sup> PD was used as reference category.
<sup>c</sup> FW levels were entered as z-scores to improve interpretability of a one-SD unit change in FW.
<sup>d</sup> Sex was coded as female = 0 and male = 1.
CI = confidence intervals; DLB = dementia with Lewy bodies; FW = free water; iRBD = isolated REM sleep behavior disorder; MoCA = Montreal Cognitive Assessment; NBM = nucleus basalis of Meynert; OR = odds ratio; PD = Parkinson's disease; pSN = posterior substantia nigra; SE = standard error.

Model 2, testing FW in the posterior SN alone, along with age and sex, was not significant (P > 0.27) (Table 2). To rule out sample composition as a confounding factor, we repeated this model using the same subset of iRBD patients included in the NBM analysis. In this restricted sample, FW in the posterior SN also did not predict conversion to DLB (P = 0.36) or PD (P = 0.75). Model 3, which included FW in both the NBM and posterior SN, along with age and sex, confirmed that NBM FW predicted phenoconversion to DLB versus remaining disease-free (B = 1.90, P < 0.001, OR = 6.69, 95% CI: 2.22 to 20.19), as well as DLB versus PD (B = 1.39, P = 0.021, OR = 4.00, 95% CI: 1.23 to 12.99) (Table 2). FW in the anterior SN (control region) was not associated with phenoconversion in any of the analyses (Table S2). Kaplan-Meier survival curves further indicated that iRBD patients with higher FW in the NBM had a significantly greater likelihood of converting to DLB over time compared to remaining disease-free (P = 0.023), with a trend also observed for conversion to PD compared to remaining disease-free (P = 0.051) (Figure 5). However, in Cox proportional hazards regression models adjusting for age and sex, only FW in the NBM remained a significant predictor of DLB conversion (HR = 8.22, 95% CI: 2.00 to 33.80, P = 0.003), while association with PD was no longer significant (HR = 1.59, 95% CI: 0.80 to 3.18, P = 0.19) (Tables S3 and S4). This indicates that increased FW content in the NBM is a specific marker of DLB phenoconversion in iRBD.

**Figure 5.**
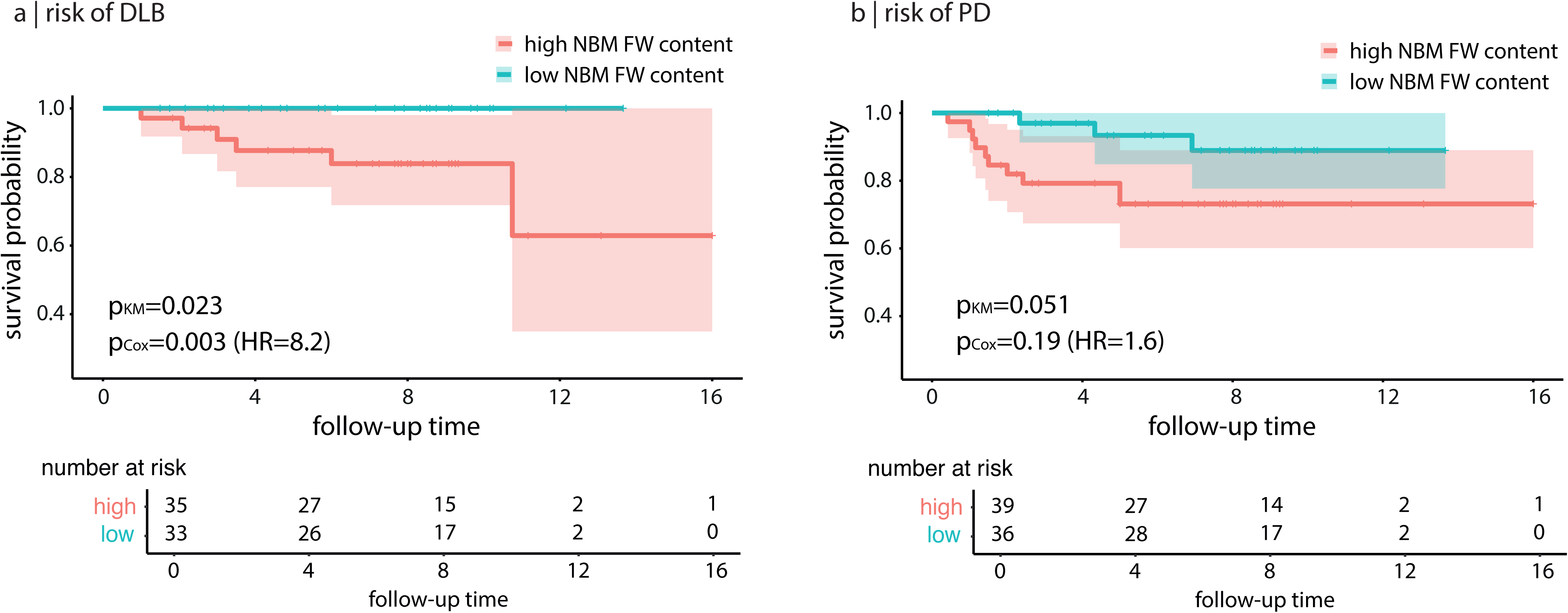
Survival analysis of disease progression trajectories in iRBD based on NBM FW content. Kaplan-Meier survival curves showing the association of FW content in the NBM to predict phenoconversion risk to **(a)** DLB and **(b)** PD, compared to remaining disease-free in iRBD. High FW content based on the median FW level is represented in red and low FW content is represented in blue. Shaded areas indicate the 95% confidence intervals for each group. FW = free water; iRBD = isolated REM sleep behavior disorder; NBM = nucleus basalis of Meynert; pSN = posterior substantia nigra.

#### NBM free water predicts conversion independently of atrophy

Next, we investigated whether FW in the NBM provided additional predictive value over structural atrophy for identifying iRBD patients at risk of phenoconversion. Mean NBM volume (normalized for head size) was significantly lower in iRBD patients (1.95 x 10^-4^, SD: 1.80 x 10^-5^) compared to controls (2.02 x 10^-4^, SD: 1.56 x 10^-5^), with a mean difference of 6.62 x 10^-6^ (95% CI: 3.36 x 10^-6^ to 9.88 x 10^-6^, P < 0.001). Among iRBD patients, converters had lower NBM volume (1.87 x 10^-4^, SD: 1.60 x 10^-5^) than non-converters (1.98 x 10^-4^, SD: 1.84 x 10^-5^), with a mean difference of 1.11 x 10^-5^ (95% CI: 5.93 x 10^-6^ to 1.62 x 10^-5^, P < 0.001) (Table 1). To test whether NBM volume could predict differential phenoconversion in iRBD, we ran a multinomial regression model adjusting for age and sex. Lower NBM volume was associated with increased odds of converting to DLB compared to remaining disease-free (B = -0.77, P = 0.044, OR = 0.46, 95% CI: 0.22 to 0.98), but not to PD (B = -0.35, P = 0.15), and did not distinguish DLB from PD (B = -0.42, P = 0.30) (Table 3). However, when both NBM FW and volume were entered into the same model, only FW in the NBM was a significant predictor of DLB conversion (B = 2.01, P = 0.035, OR = 7.45, 95% CI = 1.15 to 48.24), while NBM volume was no longer significant (B = -0.73, P = 0.16) (Table 3). These findings demonstrate that FW in the NBM is a more sensitive and specific marker of phenoconversion to DLB than structural atrophy.

**Table 3.**
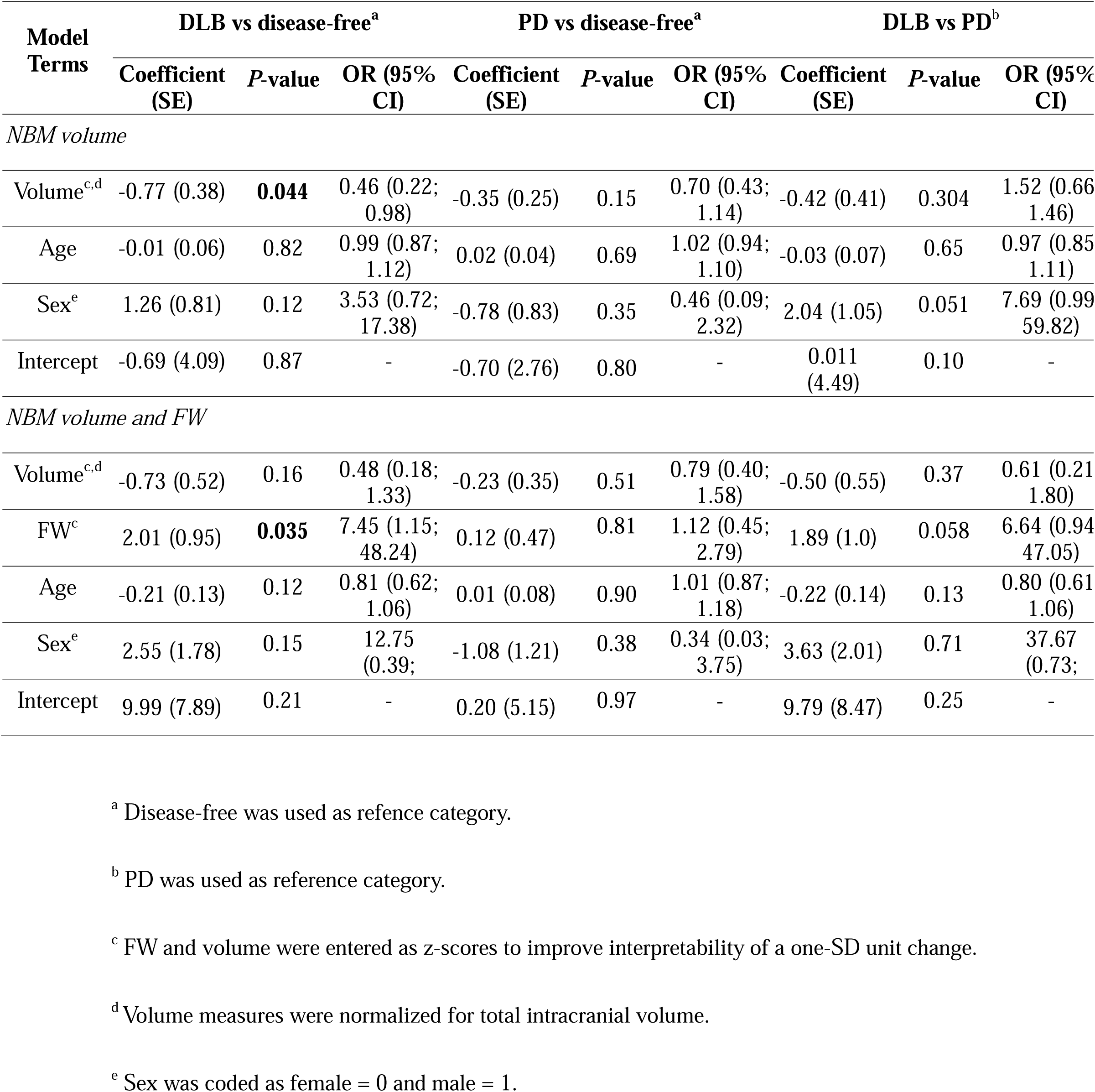
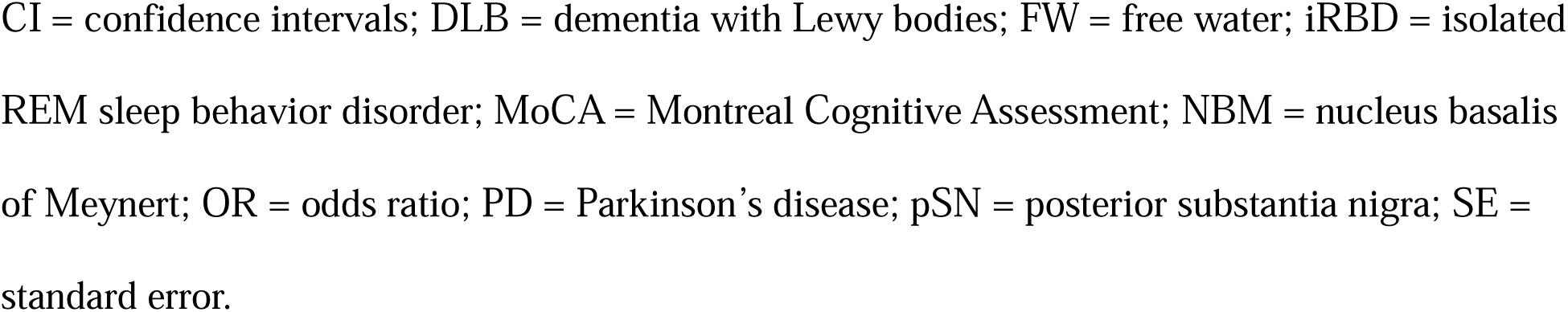
Logistic regression models of NBM volume predicting phenoconversion in iRBD.

| Model Terms | DLB vs disease-free<sup>a</sup> |  |  | PD vs disease-free<sup>a</sup> |  |  | DLB vs PD<sup>b</sup> |  |  |
| --- | --- | --- | --- | --- | --- | --- | --- | --- | --- |
|  | Coefficient (SE) | P-value | OR (95% CI) | Coefficient (SE) | P-value | OR (95% CI) | Coefficient (SE) | P-value | OR (95% CI) |
| *NBM volume* |  |  |  |  |  |  |  |  |  |
| Volume<sup>c,d</sup> | -0.77 (0.38) | **0.044** | 0.46 (0.22; 0.98) | -0.35 (0.25) | 0.15 | 0.70 (0.43; 1.14) | -0.42 (0.41) | 0.304 | 1.52 (0.66 1.46) |
| Age | -0.01 (0.06) | 0.82 | 0.99 (0.87; 1.12) | 0.02 (0.04) | 0.69 | 1.02 (0.94; 1.10) | -0.03 (0.07) | 0.65 | 0.97 (0.85 1.11) |
| Sex<sup>e</sup> | 1.26 (0.81) | 0.12 | 3.53 (0.72; 17.38) | -0.78 (0.83) | 0.35 | 0.46 (0.09; 2.32) | 2.04 (1.05) | 0.051 | 7.69 (0.99 59.82) |
| Intercept | -0.69 (4.09) | 0.87 | - | -0.70 (2.76) | 0.80 | - | 0.011 (4.49) | 0.10 | - |
| *NBM volume and FW* |  |  |  |  |  |  |  |  |  |
| Volume<sup>c,d</sup> | -0.73 (0.52) | 0.16 | 0.48 (0.18; 1.33) | -0.23 (0.35) | 0.51 | 0.79 (0.40; 1.58) | -0.50 (0.55) | 0.37 | 0.61 (0.21 1.80) |
| FW<sup>c</sup> | 2.01 (0.95) | **0.035** | 7.45 (1.15; 48.24) | 0.12 (0.47) | 0.81 | 1.12 (0.45; 2.79) | 1.89 (1.0) | 0.058 | 6.64 (0.94 47.05) |
| Age | -0.21 (0.13) | 0.12 | 0.81 (0.62; 1.06) | 0.01 (0.08) | 0.90 | 1.01 (0.87; 1.18) | -0.22 (0.14) | 0.13 | 0.80 (0.61 1.06) |
| Sex<sup>e</sup> | 2.55 (1.78) | 0.15 | 12.75 (0.39; -) | -1.08 (1.21) | 0.38 | 0.34 (0.03; 3.75) | 3.63 (2.01) | 0.71 | 37.67 (0.73; -) |
| Intercept | 9.99 (7.89) | 0.21 | - | 0.20 (5.15) | 0.97 | - | 9.79 (8.47) | 0.25 | - |

<sup>a</sup> Disease-free was used as reference category.
<sup>b</sup> PD was used as reference category.
<sup>c</sup> FW and volume were entered as z-scores to improve interpretability of a one-SD unit change.
<sup>d</sup> Volume measures were normalized for total intracranial volume.
<sup>e</sup> Sex was coded as female = 0 and male = 1.
CI = confidence intervals; DLB = dementia with Lewy bodies; FW = free water; iRBD = isolated REM sleep behavior disorder; MoCA = Montreal Cognitive Assessment; NBM = nucleus basalis of Meynert; OR = odds ratio; PD = Parkinson's disease; pSN = posterior substantia nigra; SE = standard error.

## Discussion

In this study, we leveraged the largest collection of diffusion MRI scans from vPSG-confirmed iRBD patients and healthy controls to investigate whether FW content in the NBM and posterior SN predict phenoconversion trajectories in iRBD. Our results demonstrate that changes in FW within these regions are associated with disease progression towards synucleinopathies. While no significant group differences were observed between iRBD patients and controls in either the NBM and posterior SN overall, stratification by phenoconversion status revealed that increased FW content in both regions was present among converters. Importantly, increased FW in the NBM was selectively associated with conversion to DLB, rather than PD or remaining disease-free, indicating it may serve as an early marker of DLB-specific progression. Although NBM volume was also significantly reduced in iRBD patients and in converters, only FW remained a significant predictor of DLB conversion when both measures were entered into the same model. This suggests that FW in the NBM captures disease-relevant microstructural changes beyond atrophy, offering a more sensitive biomarker of phenoconversion risk in iRBD.

Several studies have shown that increases in FW occur in overt synucleinopathies, reflecting neuroinflammation, gliosis, axonal damage, and cell loss (54). In PD, FW increases in the posterior SN and basal forebrain have been associated with motor features, attentional deficits, and memory impairments (20, 28-30). In DLB, FW increases in the NBM have been associated with cholinergic dysfunction and cognitive decline (35). Importantly, these FW changes can be observed even in the earliest stages of disease, with studies reporting increases within the first year of diagnosis in *de novo* PD patients (20, 55). In iRBD, a prodromal synucleinopathy where over 90% of patients progress to either DLB (43.5%) or PD (56.5%) (1, 2, 56), FW imaging has yielded mixed findings. While two studies reported increased FW in the posterior SN and NBM (28, 31), another found no significant FW differences in the posterior SN between iRBD patients and controls (36). In our study, we similarly found no group-level FW differences in the NBM or posterior SN when comparing all iRBD patients to controls. However, when stratifying iRBD patients based on their phenoconversion status, FW content in both the NBM and posterior SN significantly differed between groups. This indicates that FW changes are not associated with the mere presence of iRBD but rather with its progression, supporting the notion that FW is better suited as a prognostic marker than a diagnostic one. These FW differences parallel previous neuroimaging findings in iRBD, including basal forebrain atrophy (57-59), reduced neuromelanin signal in the SN (60), and decreased nigrostriatal dopaminergic function (61).

Importantly, we observed that FW increases in the NBM were specifically associated with disease progression toward DLB. Higher FW content in the NBM consistently predicted conversion to DLB, rather than PD or stable disease, even after adjusting for age and sex. Even when considering FW in the posterior SN as a covariate, odds ratios indicated that each standard deviation increase in FW was associated with a 7-fold higher risk of developing DLB compared to remaining disease-free in iRBD, and a 4-fold higher risk of developing DLB compared to developing PD, underscoring the strong predictive utility of NBM FW for DLB conversion. These findings align with recent multimodal brain-clinical models, such as a multivariate SPECT perfusion signature implicating the basal forebrain in predicting DLB versus PD trajectories in iRBD (10). Similarly, basal forebrain atrophy in iRBD has been reported (9, 62) and linked to cognitive impairment (57), and reduced cortical acetylcholinesterase activity has been associated with microglial activation in the SN (63). Longitudinal data further suggest that cholinergic dysfunction in iRBD worsens over time (64), reinforcing the vulnerability of the cholinergic system in prodromal DLB. Our results are also in line with studies showing increased FW in the NBM of patients with DLB (35). Moreover, NBM atrophy has been observed in iRBD patients with mild cognitive impairment (59), a condition considered a strong risk factor for DLB phenoconversion (65). Importantly, in this study, we demonstrate that despite showing NBM atrophy, only FW in the NBM is retained as a significant predictor of the development of DLB in iRBD. This extends the literature by showing that microstructural changes in the NBM, as indexed by FW, can serve as an early and specific marker of conversion to DLB in iRBD. That FW content in the posterior SN did not distinguish between DLB and PD converters may reflect the overlap in parkinsonian motor features shared by both disorders, which are less useful for predicting differential clinical trajectories (45).

This study has some limitations. First, although it represents the largest currently available cohort of brain MRI scans from vPSG-confirmed iRBD patients and demonstrates the feasibility of using FW as a biomarker across sites with heterogeneous imaging protocols, the number of patients who had phenoconverted to PD or DLB remains limited. Larger samples of converters and replication in independent cohorts will be essential to strengthen the generalizability of our findings. Second, we focused on phenoconversion to DLB and PD due to the small number of patients who developed MSA (N=4) or other conditions (N=6). Future studies should aim to include a more representative sample of MSA cases to better characterize the full spectrum of phenoconversion trajectories. Third, only a single MRI time point was available for each participant, which precludes the longitudinal assessment of FW dynamics over time. Fourth, clinical phenotyping across sites was limited to common variables (MoCA and MDS-UPDRS-III) and future multicentric efforts should harmonize more comprehensive clinical and neuropsychological assessments to better understand the clinical correlates of early FW changes. Fifth, *APOE* ε4 carrier status, despite its known association with faster progression to DLB (66), was not available in this dataset and could not be considered in the analyses. Sixth, region-specific challenges in quality control, particularly in the NBM due to its proximity to CSF-filled spaces, led to a reduced sample size for this region. This highlights the need for robust and automated segmentation tools tailored to deep basal forebrain structures. Nevertheless, analyses of FW in the NBM and posterior SN conducted in the same subset of participants yielded consistent findings with those from the full sample, mitigating concerns about selection bias. Finally, this study did not assess how FW changes relate to other early neurodegeneration biomarkers (e.g., PET, neuromelanin MRI or CSF markers), which may provide additional insight into underlying pathophysiological processes.

In summary, this large, multicenter, prospective study highlights the potential of FW imaging, particularly in the NBM, as a non-invasive biomarker for identifying iRBD patients at increased risk of phenoconversion to DLB. These findings support the integration of FW measures into future prognostic frameworks for prodromal synucleinopathies.

## Supporting information

Supplementary Manuscript

## Data Availability

The data for this study were sourced from multiple collaborating centers, each retaining ownership of their respective datasets. The principal investigator had authorized access to all the data required for the analyses conducted. However, data accessibility and sharing are governed by the local policies and restrictions of each participating center. Therefore, data availability is restricted, and access requests should be directed to the respective institutions, following their specific data access and sharing guidelines.

## Abbreviations

DLB: Dementia with Lewy Bodies
FW: free water
iRBD: isolated REM sleep behavior disorder
MDS: Movement Disorder Society
MoCA: Montreal Cognitive Assessment
MSA: Multiple System Atrophy
NBM: nucleus basalis of Meynert
PD: Parkinson’s Disease
SN: substantia nigra
UPDRS-III: Unified Parkinson’s Disease Rating Scale, motor scale
v-PSG: video-Polysomnography

## 6.0 Acknowledgements

C.H. holds scholarships from the Canadian Institutes of Health Research, the Quebec Bio-Imaging Network and Parkinson’s Foundation. R.B.P. reports grants from the Canadian Institute of Health Research, the Michael J. Fox Foundation, the Webster Foundation, Roche, and the National Institute of Health as well as personal fees from Takeda, Biogen, Abbvie, Curasen, Lilly, Novartis, Eisai, Paladin, Merck, Korro, Vaxxinity, Bristol Myers Squibb, and the International Parkinson and Movement Disorders Society, all outside the submitted work. S.R. holds a research scholar award from the Fonds de recherche du Québec – Santé.

C.H contributed in data analysis and draft writing; V.D contributed in data analysis and draft writing; V.A contributed in draft analysis and manuscript revision; M.F contributed in draft analysis and manuscript revision; A.P.B contributed in draft analysis and manuscript revision; C.T contributed in draft analysis and manuscript revision; A.B contributed in draft analysis and manuscript revision; M.D contributed in draft analysis and manuscript revision; A.V contributed in draft analysis and manuscript revision; J.F.G contributed in data collection and manuscript revision; R.B.P contributed in data collection and manuscript revision; P.D contributed in data collection and manuscript revision; S.M contributed in data collection and manuscript revision; Z.V contributed in data collection and manuscript revision; J.K contributed in data collection and manuscript revision; M.T.H contributed in data collection and manuscript revision; S.L contributed in data collection and manuscript revision; I.A contributed in data collection and manuscript revision; M.V contributed in data collection and manuscript revision; J.C.C contributed in data collection and manuscript revision; S.R contributed in data analysis, draft writing and study supervision.

## 6.1 Declaration of interests

All authors declare no conflict of interest.

## 6.2 Funding Sources

This work was supported by funding grants awarded to Shady Rahayel from Parkinson Canada (PPG-2023-0000000122) and Alzheimer Society Canada (#0000000082).

The work performed in Paris was funded by grants from the Programme d’investissements d’avenir (ANR-10-IAIHU-06), the Paris Institute of Neurosciences – IHU (IAIHU-06), the Agence Nationale de la Recherche (ANR-11-INBS-0006), Électricité de France (Fondation d’Entreprise

EDF), Biogen Inc., the Fondation Thérèse et René Planiol, the Fonds Saint-Michel; by unrestricted support for research on Parkinson’s disease from Energipole (M. Mallart) and Société Française de Médecine Esthétique (M. Legrand); and by a grant from the Institut de France to Isabelle Arnulf (for the Alice Study).

The work performed in Montreal was supported by the Canadian Institutes of Health Research, the Fonds de recherche du Québec – Santé, and the W. Garfield Weston Foundation. Jean-François Gagnon reports grants from the Fonds de recherche du Québec – Santé, the Canadian Institutes of Health Research, the W. Garfield Weston Foundation, and the National Institutes of Health / National Institute on Aging. He holds a Canada Research Chair in Cognitive Decline in Pathological Aging. Ronald B. Postuma reports grants and personal fees from the Fonds de recherche du Québec–Santé, the Canadian Institutes of Health Research, the Parkinson Society of Canada, the W. Garfield Weston Foundation, the Michael J. Fox Foundation for Parkinson’s Research, the R. Howard Webster Foundation, and the National Institutes of Health.

The Oxford Discovery cohort was funded by Parkinson’s UK (J-2101) and the National Institute for Health Research (NIHR) Oxford Biomedical Research Centre (BRC).

The work performed in Prague was funded by the Czech Health Research Council grant NU21-04-00535 and by project nr. LX22NPO5107 (MEYS): Financed by European Union – Next Generation EU.

The Parkinson’s Progression Markers Initiative (PPMI)—a public-private partnership—is funded by the Michael J. Fox Foundation for Parkinson’s Research and funding partners, including 4D Pharma, AbbVie Inc., AcureX Therapeutics, Allergan, Amathus Therapeutics, Aligning Science Across Parkinson’s (ASAP), Avid Radiopharmaceuticals, Bial Biotech, Biogen, BioLegend, Bristol Myers Squibb, Calico Life Sciences LLC, Celgene Corporation, DaCapo Brainscience, Denali Therapeutics, The Edmond J. Safra Foundation, Eli Lilly and Company, GE Healthcare, GlaxoSmithKline, Golub Capital, Handl Therapeutics, Insitro, Janssen Pharmaceuticals, Lundbeck, Merck & Co., Inc., Meso Scale Diagnostics, LLC, Neurocrine Biosciences, Pfizer Inc., Piramal Imaging, Prevail Therapeutics, F. Hoffman-La Roche Ltd and its affiliated company Genentech Inc., Sanofi Genzyme, Servier, Takeda Pharmaceutical Company, Teva Neuroscience, Inc., UCB, Vanqua Bio, Verily Life Sciences, Voyager Therapeutics, Inc., and Yumanity Therapeutics, Inc. For up-to-date information on the study, visit www.ppmi-info.org.

Role of the funding source: The funders had no role in data collection, analysis, interpretation, or writing of the manuscript. The corresponding author had full access to all study data and the final decision to submit for publication.

## 6.3 Consent Statement

All study participants provided written, informed consent.

## Appendix

List of the contributors involved in the ICEBERG Study Group:

Steering committee: Marie Vidailhet, MD, PhD, (Pitié-Salpêtrière Hospital, Paris, principal investigator of ICEBERG), Jean-Christophe Corvol, MD, PhD (Pitié-Salpêtrière Hospital, Paris, scientific lead), Isabelle Arnulf, MD, PhD (Pitié-Salpêtrière Hospital, Paris, member of the steering committee), Stéphane Lehericy, MD, PhD (Pitié-Salpêtrière Hospital, Paris, member of the steering committee);

Clinical data: Marie Vidailhet, MD, PhD, (Pitié-Salpêtrière Hospital, Paris, coordination), Graziella Mangone, MD, PhD (Pitié-Salpêtrière Hospital, Paris, co-coordination), Jean-Christophe Corvol, MD, PhD (Pitié-Salpêtrière Hospital, Paris), Isabelle Arnulf, MD, PhD (Pitié-Salpêtrière Hospital, Paris), Sara Sambin, MD (Pitié-Salpêtrière Hospital, Paris), Jonas Ihle, MD (Pitié-Salpêtrière Hospital, Paris), Caroline Weill, MD, (Pitié-Salpêtrière Hospital, Paris), David Grabli, MD, PhD (Pitié-Salpêtrière Hospital, Paris); Florence Cormier-Dequaire, MD (Pitié-Salpêtrière Hospital, Paris); Louise Laure Mariani, MD, PhD (Pitié-Salpêtrière Hospital, Paris), Bertrand Degos, MD, PhD (Avicenne Hospital, Bobigny);

Neuropsychological data: Richard Levy, MD (Pitié-Salpêtrière Hospital, Paris, coordination), Fanny Pineau, MS (Pitié-Salpêtrière Hospital, Paris, neuropsychologist), Julie Socha, MS (Pitié-Salpêtrière Hospital, Paris, neuropsychologist), Eve Benchetrit, MS (La Timone Hospital, Marseille, neuropsychologist), Virginie Czernecki, MS (Pitié-Salpêtrière Hospital, Paris, neuropsychologist), Marie-Alexandrine, MS (Pitié-Salpêtrière Hospital, Paris, neuropsychologist);

Eye movement: Sophie Rivaud-Pechoux, PhD (ICM, Paris, coordination); Elodie Hainque, MD, PhD (Pitié-Salpêtrière Hospital, Paris);

Sleep assessment: Isabelle Arnulf, MD, PhD (Pitié-Salpêtrière Hospital, Paris, coordination), Smaranda Leu Semenescu, MD (Pitié-Salpêtrière Hospital, Paris), Pauline Dodet, MD (Pitié-Salpêtrière Hospital, Paris);

Genetic data: Jean-Christophe Corvol, MD, PhD (Pitié-Salpêtrière Hospital, Paris, coordination), Graziella Mangone, MD, PhD (Pitié-Salpêtrière Hospital, Paris, co-coordination), Samir Bekadar, MS (Pitié-Salpêtrière Hospital, Paris, biostatistician), Alexis Brice, MD (ICM, Pitié-Salpêtrière Hospital, Paris), Suzanne Lesage, PhD (INSERM, ICM, Paris, genetic analyses);

Metabolomics: Fanny Mochel, MD, PhD (Pitié-Salpêtrière Hospital, Paris, coordination), Farid Ichou, PhD (ICAN, Pitié-Salpêtrière Hospital, Paris), Vincent Perlbarg, PhD, Pierre and Marie Curie University), Benoit Colsch, PhD (CEA, Saclay), Arthur Tenenhaus, PhD (Supelec, Gif-sur-Yvette, data integration);

Brain MRI data: Stéphane Lehericy, MD, PhD (Pitié-Salpêtrière Hospital, Paris, coordination), Rahul Gaurav, MS, (Pitié-Salpêtrière Hospital, Paris, data analysis), Nadya Pyatigorskaya, MD, PhD, (Pitié-Salpêtrière Hospital, Paris, data analysis); Lydia Yahia-Cherif, PhD (ICM, Paris, Biostatistics), Romain Valabregue, PhD (ICM, Paris, data analysis), Cécile Galléa, PhD (ICM, Paris);

DaTscan imaging data: Marie-Odile Habert, MCU-PH (Pitié-Salpêtrière Hospital, Paris, coordination);

Voice recording: Dijana Petrovska, PhD (Telecom Sud Paris, Evry, coordination), Laetitia Jeancolas, MS (Telecom Sud Paris, Evry);

Study management: Vanessa Brochard (Pitié-Salpêtrière Hospital, Paris, coordination), Alizé Chalançon (Pitié-Salpêtrière Hospital, Paris, Project manager), Carole Dongmo-Kenfack (Pitié-

Salpêtrière Hospital, Paris, clinical research assistant); Christelle Laganot (Pitié-Salpêtrière Hospital, Paris, clinical research assistant), Valentine Maheo (Pitié-Salpêtrière Hospital, Paris, clinical research assistant).

## Notes

### Competing Interest Statement

The authors have declared no competing interest.

### Author Declarations

Ethics committee of Hopital du Sacre-Coeur de Montreal (CIUSSS-NIM) gave ethical approval for this work. Ethics committee of McGill University gave ethical approval for this work.

