## Supplementary Manuscript for "Free water predicts dementia with Lewy bodies in isolated REM sleep behavior disorder": Supplementary Manuscript .pdf

**Table of contents:**

Acquisition parameters of diffusion- and T1-weighted brain MRI.

Figure S1. FW processing workflow.

Figure S2. FW content in control regions in iRBD patients and controls.

Figure S3. FW content in control regions in iRBD converters and non-converters.

Table S1. Cox proportional hazards regression model predicting phenoconversion risk in iRBD.

Table S2. Logistic regression models predicting progression trajectories in iRBD using aSN.

Table S3. Cox proportional hazards model for phenoconversion to DLB in iRBD.

Table S4. Cox proportional hazards model results for phenoconversion to PD in iRBD.

### Acquisition parameters of diffusion- and T1-weighted brain MRI.

The DWI scans from Montreal were acquired using a 3T Siemens TIM Trio scanner with a 12-channel head coil and echo-planar sequence with the following parameters: b-values of 0 and 700 s/mm$^2$; gradient directions of 63 and 64; 2-mm isotropic voxel size; TR=8.6-12.7 ms; TE=0.083-0.1 ms; or using a 3T Siemens PRISMA scanner with a 32-channel head coil and echo-planar sequence: b-values of 0 and 1,000 s/mm$^2$; gradient directions of 30; 2-mm isotropic voxel size; TR=6.9 ms; TE=64 ms. The Prague cohort was scanned using a 3T Siemens Skyra with a 32-channel head coil and echo-planar sequence: b-values of 0 and 1,000 s/mm$^2$; gradient directions of 29 or 30; 2-mm isotropic voxel size; TR=10.5 ms; TE=93 ms. The Oxford cohort was scanned using a 3T Siemens Trio scanner with a 12-channel head coil and echo-planar sequence: b-values of 0 and 1,000 s/mm$^2$; gradient directions of 60; 2-mm isotropic voxel size; TR=9.3 ms; TE=94 ms. The Paris cohort was scanned using a 3T Siemens TIM Trio scanner with a 12-channel head coil and echo-planar imaging: b-values of 0 and 700 s/mm$^2$; gradient directions of 21; 1.72-mm isotropic voxel size; TR=14 ms; TE=101 ms; or using a 3T PRISMA Fit scanner with a 64-channel head coil and echo-planar imaging: b-values of 0 and 700 s/mm$^2$; gradient directions of 32; 1.7-mm isotropic voxel size; TR=10.4 ms; TE=59 ms.

The acquisition parameters for the T1-weighted MRI scans from the Montreal cohort were acquired with an MPRAGE sequence with the following parameters: TR=2,300 ms; TE=2.91 ms; flip angle=9°; and 1-mm isotropic voxel size; or with TR=2,300 ms; TE=2.98 ms; flip angle=9°; and 1-mm isotropic voxel size. The Prague cohort used an MPRAGE sequence with TR=2,200 ms; TE=2.4 ms; flip angle=8°; and 1-mm isotropic voxel size. The Oxford cohort was scanned using an MPRAGE sequence with TR=2,040 ms; TE=4.7 ms; flip angle=8°; and 1-mm isotropic voxel size. The Paris cohort used an MPRAGE sequence with TR=2,300 ms; TE=4.18 ms; flip angle=9°; and 1-mm isotropic voxel size; or with an MP2RAGE sequence: TR=5,000 ms, TE=2.98 ms, flip angles=4° and 5°; GRAPPA=3, and 1-mm isotropic voxel size. Refer to www.ppmi-info.org for the T1- and diffusion-weighted images from the PPMI study (15).

**Figure S1. FW processing workflow.**

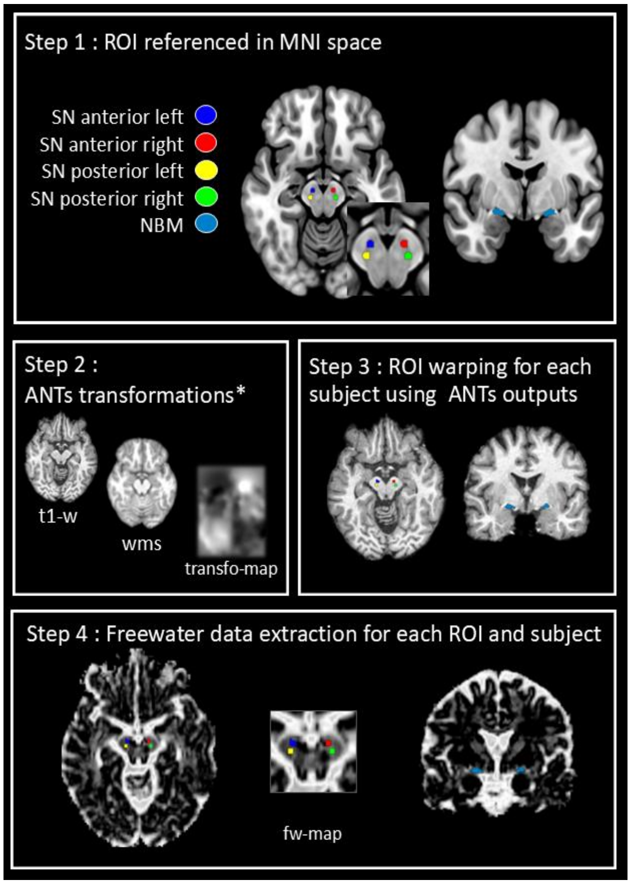


Workflow for extracting FW values from ROIs. ROIs, including basal forebrain and posterior and anterior SN, were defined in MNI space. ANTs transformations were applied to align individual T1-weighted images to MNI space. ROIs were warped into each subject's native space using the transformation outputs. FW values were extracted from FW maps for each ROI in native space.

ANTs = Advanced Normalization Tools; NBM = nucleus basalis of Meynert; FW = free water; fw-map = free water map; MNI = Montreal Neurological Institute; ROI = region of interest; SN = substantia nigra; t1-w = T1-weighted Image; transfo-map = transformation map; wms = white matter segmentation.

**Figure S2. FW content in aSN in iRBD patients and controls.**

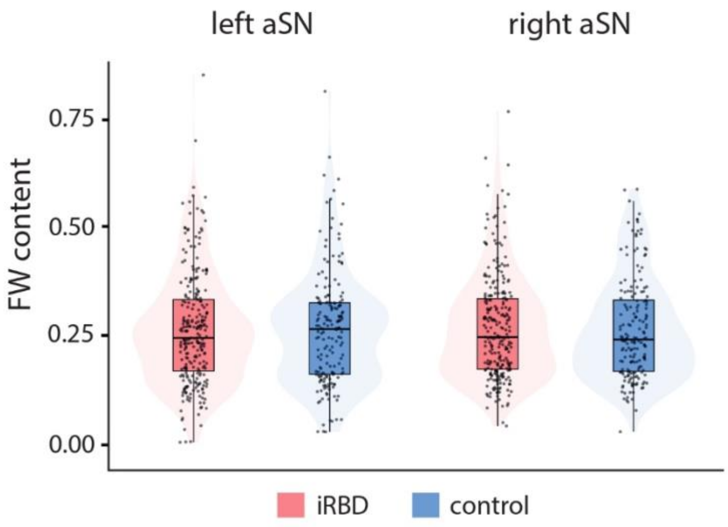


Box plots and violin plots showing the distribution of FW content in the aSN, used as a control region, for iRBD patients (red) and healthy controls (blue). FW content is presented after harmonization for scanner effects, with values standardized to positive values for visualization purposes. FW content in the aSN was not significantly different in iRBD patients compared to healthy controls.

aSN = anterior substantia nigra; FW = free water; iRBD = isolated REM sleep behavior disorder.

**Figure S3. FW content in aSN in iRBD converters and non-converters.**

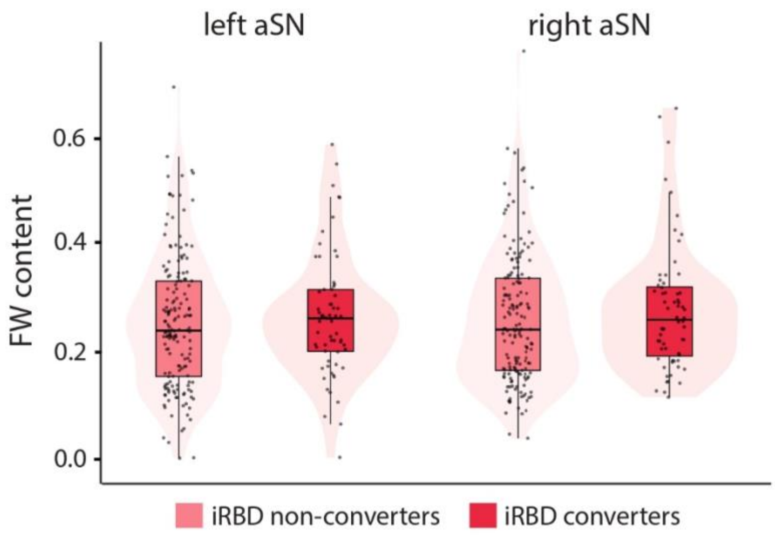


Box plots and violin plots showing the distribution of FW content in the aSN control region for iRBD converters and non-converters. FW content is presented after harmonization for scanner effects, with values standardized to positive values for visualization purposes. FW content in the aSN was not significantly different in iRBD converters compared to non-converters.

aSN = anterior substantia nigra; FW = free water; iRBD = isolated REM sleep behavior disorder; NBM = nucleus basalis of Meynert.

**Table S1. Cox regression models predicting phenoconversion risk in iRBD.**

| Model Terms | Coefficient | Standard error | Hazard ratio | Lower 95% CI | Upper 95% CI | P-value |
|---|---|---|---|---|---|---|
| ***Model 1: NBM FW*** | | | | | | |
| Sex | -0.86 | 0.70 | 0.42 | 0.11 | 1.67 | 0.22 |
| Age | -0.06 | 0.04 | 0.94 | 0.87 | 1.02 | 0.11 |
| NBM FW[a] | 0.85 | 0.28 | 2.35 | 1.36 | 4.05 | **0.0021** |
| ***Model 2: pSN FW*** | | | | | | |
| Sex | -0.06 | 0.44 | 0.95 | 0.40 | 2.23 | 0.90 |
| Age | 0.01 | 0.02 | 1.01 | 0.97 | 1.06 | 0.58 |
| pSN FW[a] | 0.29 | 0.14 | 1.33 | 1.00 | 1.77 | **0.048** |
| ***Model 3: NBM and pSN FW*** | | | | | | |
| Sex | -0.90 | 0.71 | 0.40 | 0.10 | 1.61 | 0.20 |
| Age | -0.06 | 0.04 | 0.94 | 0.87 | 1.02 | 0.12 |
| NBM FW[a] | 0.83 | 0.28 | 2.30 | 1.33 | 3.98 | **0.0028** |
| pSN FW[a] | 0.11 | 0.21 | 1.12 | 0.74 | 1.69 | 0.58 |

The model's likelihood ratio $\chi^2$ value was 12.35 (P = 0.01). Bold values represent significant predictors in the model.

[a] FW content was z-scored before being entered into the model; coefficients therefore reflect a one-SD unit change in the predictor.

CI = confidence interval; FW = free water; iRBD = isolated REM sleep behavior disorder; NBM = nucleus basalis of Meynert; pSN = posterior SN; SD = standard deviation.

**Table S2. Logistic regression models predicting progression trajectories in iRBD using aSN.**

| Model Terms | DLB vs disease-free[a] | | | PD vs disease-free[a] | | | DLB vs PD[b] | | |
|---|---|---|---|---|---|---|---|---|---|
| | **Estimate (SE)** | ***P*-value** | **OR (95% CI)** | **Estimate (SE)** | ***P*-value** | **OR (95% CI)** | **Estimate (SE)** | ***P*-value** | **OR (95% CI)** |
| FW[c] | 0.19 (0.24) | 0.45 | 1.21 (0.75; 1.95) | 0.01 (0.18) | 0.96 | 1.01 (0.71; 1.44) | 0.18 (0.29) | 0.54 | 1.19 (0.68; 2.10) |
| Age | 0.01 (0.04) | 0.87 | 1.01 (0.93; 1.09) | 0.04 (0.03) | 0.17 | 1.04 (0.98; 1.10) | -0.03 (0.05) | 0.50 | 0.97 (0.88; 1.06) |
| Sex[d] | 0.41 (0.68) | 0.54 | 1.51 (0.40; 5.66) | -0.99 (0.19) | 0.19 | 0.37 (0.08; 1.64) | 1.40 (0.97) | 0.15 | 4.05 (0.60; 27.14) |
| Intercept | -3.10 (2.72) | 0.26 | - | -4.22 (1.94) | **0.030** | **-** | 1.12 (3.19) | 0.73 | - |

[a] The disease-free group was used as reference category.

[b] PD was used as reference category.

[c] FW content was z-scored to improve interpretability of a one-SD change in FW.

[d] Sex was coded as 0=female and 1=male.

aSN = anterior substantia nigra; CI = confidence intervals; DLB = dementia with Lewy bodies; FW = free water; iRBD = isolated REM sleep behavior disorder; OR = odds ratio; PD = Parkinson's disease; SE = standard error.

**Table S3. Cox regression model of NBM FW for phenoconversion to DLB in iRBD.**

| Variable | Coefficient | Standard error | Hazard ratio | Lower 95% CI | Upper 95% CI | P-value |
|---|---|---|---|---|---|---|
| Sex[a] | -3.05 | 1.61 | 0.05 | 0.002 | 1.11 | 0.06 |
| Age | -0.19 | 0.10 | 0.83 | 0.69 | 1.01 | 0.06 |
| NBM FW[b] | 2.11 | 0.72 | 8.22 | 2.00 | 33.80 | **0.003** |

The model's likelihood ratio $\chi^2$ value was 11.13 (P = 0.01). Bold values represent significant predictors in the model.

[a] Sex was coded as 0=female and 1=male.

[b] FW content was z-scored to improve interpretability of a one-SD change in FW.

CI = confidence intervals; DLB = dementia with Lewy bodies; FW = free water; iRBD = isolated REM sleep behavior disorder; NBM = nucleus basalis of Meynert; SD = standard deviation.

**Table S4. Cox regression model of NBM FW for phenoconversion to PD in iRBD.**

| Variable | Coefficient | Standard error | Hazard ratio | Lower 95% CI | Upper 95% CI | P-value |
|---|---|---|---|---|---|---|
| Sex[a] | 0.10 | 1.11 | 1.10 | 0.13 | 9.64 | 0.93 |
| Age | -0.02 | 0.05 | 0.98 | 0.89 | 1.09 | 0.75 |
| NBM FW[b] | 0.47 | 0.35 | 1.59 | 0.80 | 3.18 | 0.19 |

The model's likelihood ratio $\chi^2$ value was 2.29 and non-significant (P=0.50).

[a] Sex was coded as 0=female and 1=male.
[b] FW content was z-scored to improve interpretability of a one-SD change in FW.

CI = confidence intervals; FW = free water; iRBD = isolated REM sleep behavior disorder; NBM = nucleus basalis of Meynert; PD = Parkinson's disease; SD = standard deviation.
